# Sex differences in symptomatology and immune profiles of Long COVID

**DOI:** 10.1101/2024.02.29.24303568

**Authors:** Julio Silva, Takehiro Takahashi, Jamie Wood, Peiwen Lu, Alexandra Tabachnikova, Jeff R. Gehlhausen, Kerrie Greene, Bornali Bhattacharjee, Valter Silva Monteiro, Carolina Lucas, Rahul M. Dhodapkar, Laura Tabacof, Mario Peña-Hernandez, Kathy Kamath, Tianyang Mao, Dayna Mccarthy, Ruslan Medzhitov, David van Dijk, Harlan M. Krumholz, Leying Guan, David Putrino, Akiko Iwasaki

## Abstract

Strong sex differences in the frequencies and manifestations of Long COVID (LC) have been reported with females significantly more likely than males to present with LC after acute SARS-CoV-2 infection^1–7^. However, whether immunological traits underlying LC differ between sexes, and whether such differences explain the differential manifestations of LC symptomology is currently unknown. Here, we performed sex-based multi-dimensional immune-endocrine profiling of 165 individuals^8^ with and without LC in an exploratory, cross-sectional study to identify key immunological traits underlying biological sex differences in LC. We found that female and male participants with LC experienced different sets of symptoms, and distinct patterns of organ system involvement, with female participants suffering from a higher symptom burden. Machine learning approaches identified differential sets of immune features that characterized LC in females and males. Males with LC had decreased frequencies of monocyte and DC populations, elevated NK cells, and plasma cytokines including IL-8 and TGF-β-family members. Females with LC had increased frequencies of exhausted T cells, cytokine-secreting T cells, higher antibody reactivity to latent herpes viruses including EBV, HSV-2, and CMV, and lower testosterone levels than their control female counterparts. Testosterone levels were significantly associated with lower symptom burden in LC participants over sex designation. These findings suggest distinct immunological processes of LC in females and males and illuminate the crucial role of immune-endocrine dysregulation in sex-specific pathology.

## Introduction

Long COVID (LC) has emerged as a growing consequence and public health crisis of the ongoing COVID-19 pandemic, now in its fourth year. LC manifests as a multifaceted constellation of diverse and often debilitating symptoms, affecting multiple organ systems and varying greatly in severity. An estimated 65 million people worldwide live with LC^9,10^. Understanding the pathobiology of this disease state is a crucial initial step for developing therapies.

LC shares many traits with other post-acute infection syndromes (PAIS) which have been described following many infectious illnesses^5^, including fatigue, post-exertional malaise, pain, and neurocognitive symptoms, with many symptoms showing a strong female sex bias^11^. Unlike acute COVID-19, where males face higher severity and mortality^12–15^, LC has shown a strong repeated female bias in both the rate of diagnosis and symptomology, with females experiencing higher rates of headaches and neurological conditions, and males showing endocrine dysfunction^1,3^.

Several non-mutually exclusive hypotheses exist to explain LC, each with varying levels of support. These potential causes include the persistence of a viral reservoir, induction of cellular and humoral autoimmunity, reactivation of latent herpesviruses (such as EBV, VZV and HHV-6), microbiota dysregulation, vascular endothelial damage, persistent immune dysregulation, and dysfunctional neurological signaling^5^. Work remains to develop and understanding how each of these may contribute, together or in isolation, to the constellation of LC manifestations being reported clinically and in turn how they may manifest across sex.

As with acute COVID-19, where biological sex differences were associated with distinct immune responses^16^ and health outcomes^12^, the contribution of biological sex to LC prevalence and symptom manifestation has yet to be explained. Understanding the biological associations that may help explain the strong female bias in this condition may further help us understand LC and identify possible therapeutic strategies. To investigate the sex differences in the immunological properties of LC, we performed sex-based multi-dimensional immune-endocrine profiling of 165 individuals with or without LC enrolled through the MY-LC study in an exploratory, cross-sectional study^8^. We employed machine learning techniques to identify unique immune signatures across sexes and to associate these with the unique symptom profiles in LC.

## Results

### Overview of the study participants

185 participants were initially enrolled at Mount Sinai Hospital, in New York City, New York, composed of 101 individuals with LC (69 females, 32 males), and 82 controls (58 females, 24 males)^8^. Post-enrollment, a comprehensive review of electronic medical records led to the identification of 20 individuals with potential confounding factors: outlier conditions (n=7), oral steroid use (n=4), mismatched data (n=3), and missing data (n=6). For our primary analysis, these 20 individuals were excluded from the primary dataset, leading to a refined cohort of 165 participants.

#### Demographics and Comorbidities

To explore biological sex differences in LC we categorized our cohort into four major sex-based groups: control females (n= 53), consisting of 27 uninfected healthy controls, and 26 convalescent control individuals; control males (n=23) consisting of 12 uninfected and 11 convalescent controls; females with LC (n=58); and males with LC (n=31) (Figure 1a). Sex designation was obtained from healthcare records, corresponding to the sex assigned at birth for all individuals. A breakdown of sex-designation and self-identified gender designations is provided in Extended Data Table 1. To understand the comparability of our cohort we measured the distribution of demographic factors including age, Body Mass Index (BMI), and race/ethnicity across the four sex-based groups. Sex-based groups showed no significant difference in their age distributions, with most subjects falling into the 30-50 and 50-65 age ranges for both participants with LC and controls, though a marginally older skew amongst individuals with LC was appreciated (Figure 1b, and Extended Data Table 1). Similarly, there was no significant difference in BMI or race/ethnicity distributions across sex-based groups (Extended Data Table 1, and Extended Data Figure 1a). Vaccination status was significantly different across groups, with females with LC showing lower rates of vaccination (Extended Data Table 1).

**Figure 1:**
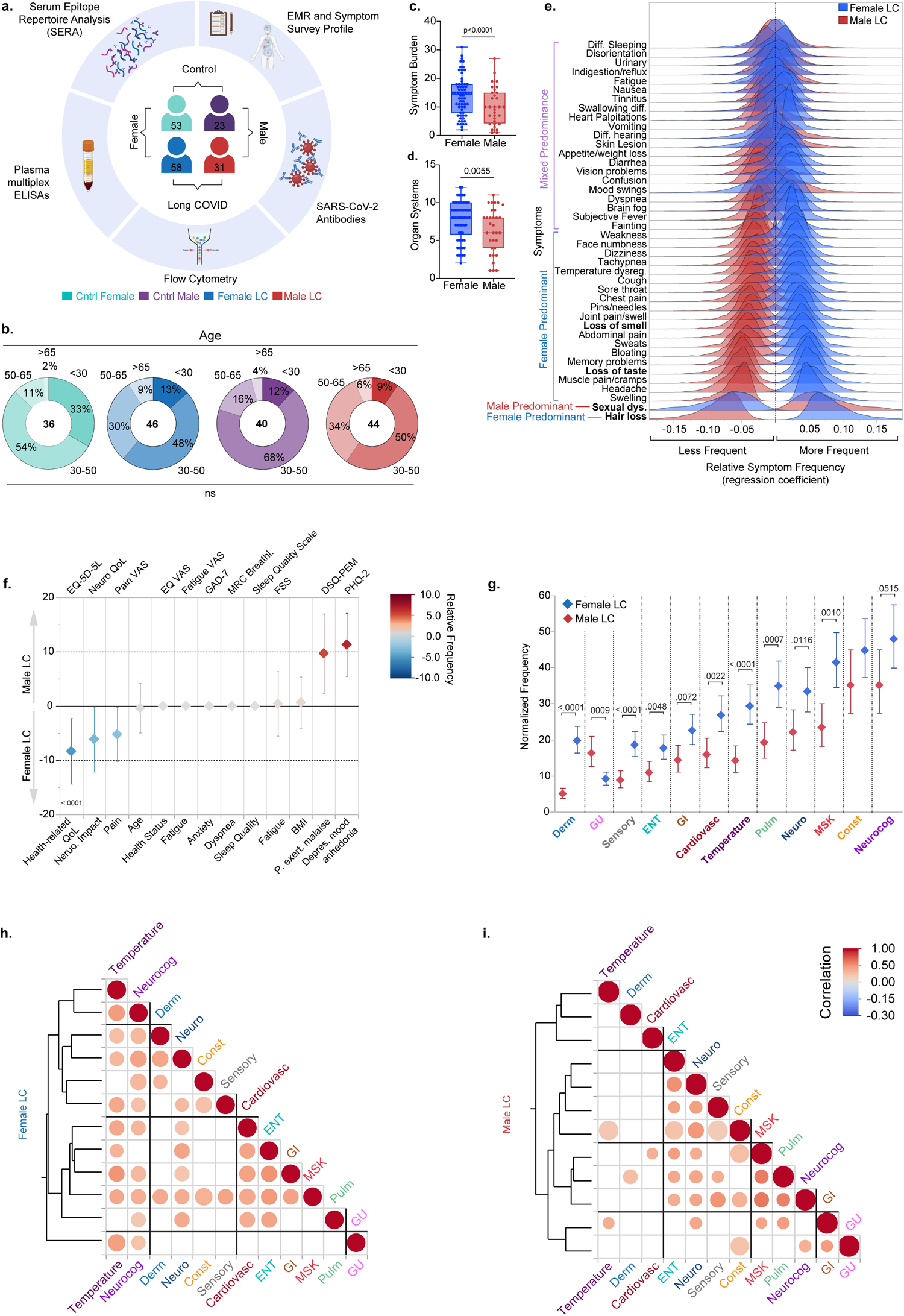
Long COVID is experienced differently by sex. (a) schematic of MYLC study. (b) age distribution across the cohort for biologically relevant age groups. Age groups frequencies were compared using binomial logistic regression using Generalized Linear Models (GLM) with odds ratios comparison of each sex and age-based group to the overall average using Analysis of Means (ANOM) with significance adjustment using the Nelson method. (c, d) comparison of counts of symptom burden and organ system involvement across sex using Poisson regression corrected for multiple comparisons using the Benjamini-Hochberg method. (e) Kernel density plots depicting the bootstrapped regression coefficients (relative symptom frequency) across sex obtained from Partial Least Squares Discriminant Analysis (PLS-DA). Symptoms are ordered by the difference in frequency across sex with least difference above and most difference below for additional details of the PLS model please see supplemental Tables. (f) Binomial Lasso regression with 5-fold cross validation comparing the relative frequencies of standardized health surveys across sex while accounting for age and BMI. Surveys were adjusted such that higher values represent worse outcomes. Whiskers represent the 95% confidence interval of each predictor. Survey names are displayed above each column with descriptions displayed below. (g) Adjusted estimated mean of normalized symptom count by organ system for females and males with Long COVID. Means were obtained using a generalized linear mixed model (GLMM) with a Poisson distribution, accounting for age and BMI. Individual participant variability was accounted for as a random effect. Graph depicts the adjusted means by sex for each organ system and whiskers represent the 95% CI of the mean. Comparisons were made across sex within each organ system. Benjamini FDR adjusted p-values are shown. (h, i) Correlational heatmap using unsupervised hierarchical clustering of Spearman correlations comparing the relationship across organ systems. Values are bootstrapped and only significant circles are shown. Size of circles is proportional to the inverse differences of the 95% confidence intervals. Abbreviations for Organ systems are as follows: Derm: Dermatological; GU: Genitourinary; Sensory: Sensory; ENT: Ear, Nose, and Throat; GI: Gastrointestinal Cardiovasc: Cardiovascular; Pulm: Pulmonary; Neuro: Neurological; MSK: Musculoskeletal; Const: Constitutional; Neurocog: Neurocognitive. Detailed descriptions and definitions of survey, organ system, and symptom terms can be found in the Data Dictionary.

We next sought to understand whether our LC participant cohort showed enrichment of comorbidities and whether these comorbidities may be sex-associated. To do so, we classified each participant’s pre-existing comorbidities into 14 major health system categories and compared the relative frequencies of each of these categories across our sex-based groups while accounting for age and BMI (Extended Data Table 1 and Extended Data Table 2). As previously reported^17,18^, there was significant enrichment of asthma in our female with LC group (p=0.0189) compared to their control female counterpart. In addition, females with LC demonstrated higher rates of gastrointestinal conditions. We also compared the frequency of sex hormonal therapies and other therapies known to cause changes in hormone levels across groups and found no significant differences.

### Symptomology of LC varies between sexes

We first measured the overall symptom burden and organ system involvement in females and males with LC, by counting the number of reported symptoms per individual and their classification of one of 12 organ systems (Extended Data Table 3). Females showed significantly higher symptom burden (p<.0001, Figure 1c) and organ system involvement (p=0.0055, Figure 1d) in our cohort compared to males with LC. This finding persisted even after accounting for age, BMI, and other comorbidities, or grouping classifications (Extended Data Figure 1b, c). Respiratory/pulmonary symptoms were also positively associated with symptom burden after adjustment. Unvaccinated individuals showed significantly higher symptom burden. Notably being on sex hormone therapy was negatively associated with symptom burden (Extended Data Figure 1b).

To explore the differences in symptom frequency between females and males with LC, we used a supervised dimensionality reduction technique, Partial Least Squares Discriminant Analysis (PLS-DA) with 5-fold cross-validation. We used PLS-DA to differentiate between females and males with LC based using the frequency of 42 common symptoms reported by both sexes. One PLS-DA component was selected for this model based on the five-fold cross-validation process. Post-analysis, each symptom’s regression coefficient, constituting the relative symptom frequency across females and males, was bootstrapped to provide a distribution reflecting how strongly and in what direction (positive or negative) each symptom was associated with females or males with LC (Figure 1e).

Our findings indicated that some LC symptoms were equally reported across males and females with LC, including commonly reported symptoms like sleep disturbance and fatigue, and less frequent symptoms such as urinary incontinence (Figure 1e and Extended Figure 1d). Notably, however, females with LC had higher overall frequencies of symptoms spanning multiple organ systems including changes in body temperature, cough, and neurological and neurocognitive symptoms such as headaches and confusion. The top distinguishing symptoms of LC status by sex were hair loss in females and sexual dysfunction in males (Figure 1e). These findings further suggested that females with LC experienced a great amount of symptom burden compared to males with LC, but the relative frequency of specific symptoms could vary across sexes. Standardized questionnaires such as the EQ-5D-5L and others are used to determine the overall health status of individuals and assess the impact that a chronic condition may have on their self-perception and their quality of life. We compared the results of these surveys across females and males with LC using binomial logistic regression with least absolute shrinkage and selection operator (LASSO) regularization, to identify only those surveys that showed the most difference between sexes. Interestingly, we noted significant differences in the severity scores for some of these surveys that distinguished females and males with LC. Females with LC reported higher pain, higher impact on their overall health, and higher neurological impact. By contrast, males with LC reported higher impacts on mood and higher rates of post-exertional malaise (Figure 1f). These differences in sex-specific reporting on standardized surveys were not seen when we compared control males to control females (Extended Data Figure 1e). We also measured the frequency of reporting organ system involvement across sexes. Consistently, females showed higher rates of reporting in many of the organ systems (Figure 1g). Finally, we aimed to explore the interconnections between the organ systems by analyzing the correlations among them across sex using unsupervised hierarchical clustering. Notably, females and males showed distinct symptom clusters. In addition to neurological and neurocognitive symptoms in both females and males with LC, temperature-related symptoms and musculoskeletal symptoms were also strongly associated with other symptom groups in females (Figure 1h). By contrast, ear nose and throat (ENT) symptoms were most associated with other symptom groups in males (Figure 1i). Taken together these findings suggest that males and females experienced a distinct LC symptom profile and pattern.

### Machine Learning identifies sex-specific LC immune signatures

To investigate sex-specific immune responses in LC, we conducted a sex-stratified analysis aimed at distinguishing between sex-predominant and sex-independent phenotypes (Figure 2). We first analyzed the immune profiles of vaccinated individuals of both control and LC groups without pre-existing autoimmune or hormone conditions or sex-hormone therapy, in males and females separately. These profiles included cytokines, hormones, cell type frequencies, antibody levels against SARS-CoV-2 as well as antibody reactivity against common latent herpesviruses including EBV, CMV, and HHV6-b, as measured by linear epitope mapping. We also included antibody cumulative linear peptide scores against infectious agents associated with chronic infection or latency and the development of PAIS or autoimmunity including reactivities to *Babesia microti, Helicobacter pylori, and Borrelia burgdorferi.* Finally, cumulative scores for common autoreactivities were also included such as the Smith Antibodies (Extended Data Figure 2). We employed PLS-DA with 5-fold cross-validation to extract LC immune signatures. For each final PLS-DA model, we subsequently constructed a variable of importance projection (VIP) by regression plot, bootstrapped for both the regression coefficient and the importance measure. This allowed us to identify the most significant features that predicted LC status for each sex separately, considering the magnitude and direction of each feature’s association with LC status (Figure 2a, and Extended Figure 2 a, b). Features with 95% confidence intervals above the threshold cutoff of 0.8 and whose regression coefficient did not pass zero, were considered most significant and important. Detailed reports for each analysis can be found in supplemental data (Supplemental Data 1).

**Figure 2:**
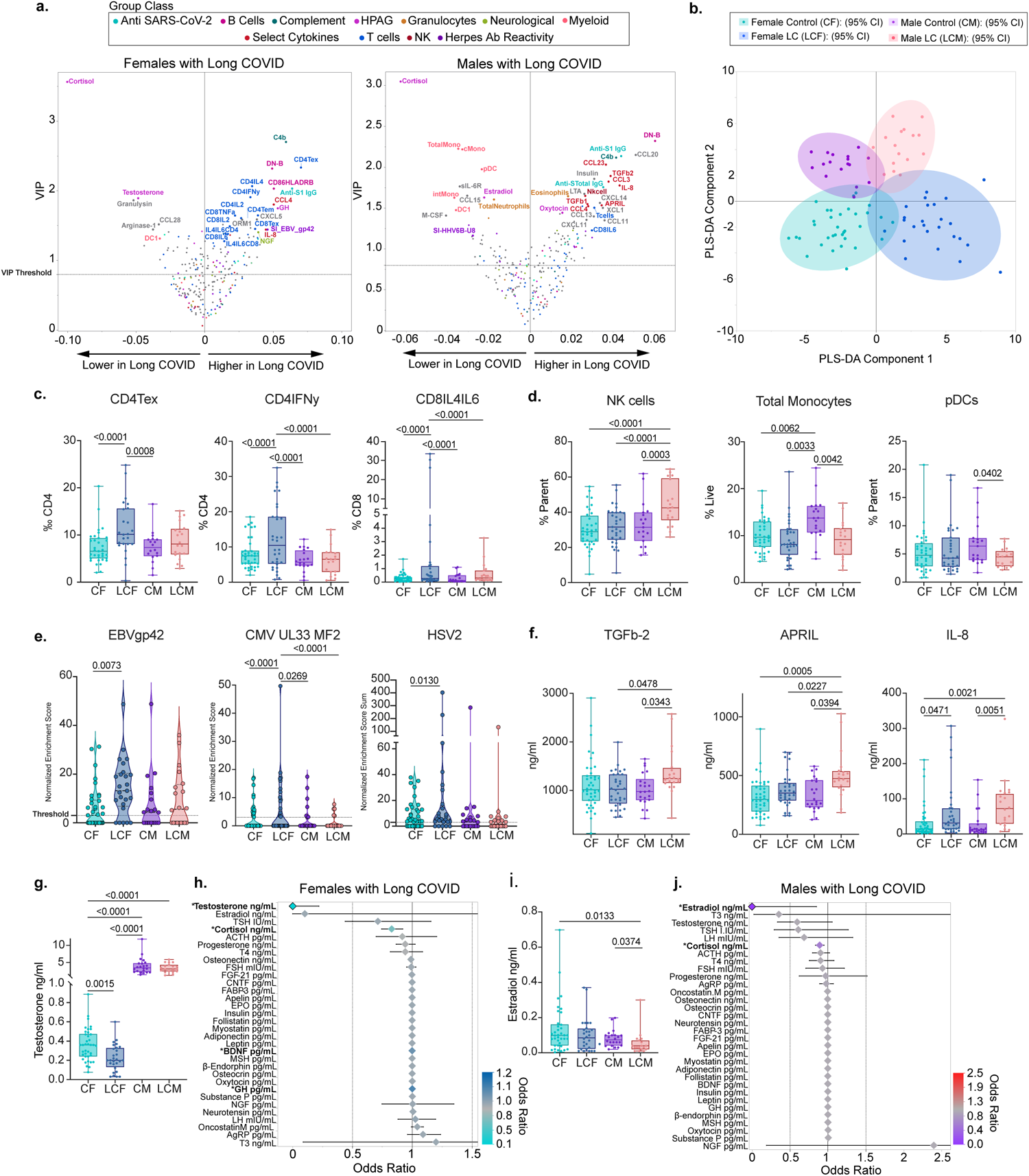
Females and males show a distinct immune profile associated with Long COVID status. (a) Sex-stratified Variable importance Projection (VIP) by regression plots for prediction of Long COVID status versus controls amongst vaccinated individuals, without pre-existing autoimmune or hormone conditions or sex-hormone therapy. Data shows Females with Long COVID versus control females (left); and males with Long COVID versus control males (right). VIP by regression Plots were derived from PLS-DA using the NIPALS algorithm with 5-fold cross validation. Each VIP score was bootstrapped to obtain 95% confidence intervals. Dotted line denotes the threshold cutoff for feature importance. Regression is denoted on the x-axis with solid line indicating the intercept. Positive values indicate positive predictors of Long COVID status and Negative values indicate negative predictors of Long COVID status. Each dot represents a unique feature included in the model (see Extended Data Figure 2, and Supplementary Tables). Color scheme denotes the classification of each feature as shown in the key. Figure annotations: exhausted CD4 and CD8T cells (CD4Tex, CD8Tex) Select Cytokines label was provided to cytokines showing a disproportionate male bias. Only features with 95% CIs above the VIP Threshold and regression 95% CI that do not pass the intercept were considered most significant and are labeled above. All features are shown in Extended Figure 2. (b) PLS-DA-derived component plot from the first 2 components of the sex-integrated PLS-DA model accounting for 27% (PLS-DA Component 1) and 24% (PLS-DA Component 2) of the variability in y respectively. Model was optimized at 6 components (cumulative pseudo R-squared: 0.84). Each Dot represents an individual. 95% confidence ellipses are shown for each sex-based group and color-coded as per the key. Additional feature analysis is shown in Extended Data Figure 3. (c) GLMs with maximum likelihood estimation (MLE), comparing the frequency of T cell types amongst vaccinated individuals across sex-based groups. Group inclusion is as in (a,b). For frequencies less than 1 (i.e. CD4 Tex) beta distribution was used with a logit model link transformation. For frequencies greater than 1, (CD4 IFN-γ, and CD8 IL4^+^/IL6^+^ double positive) negative binomial distribution was used with a Log Link transformation. (d) GLMs with MLE, comparing NK, Total Monocytes, and pDCs across sex-based groups using negative binomial regression (NK, Total Monocytes) and Poisson Regression (pDCs) with Long Link transformation. Significance was derived from the estimate of each factor in the model. (e) GLMs with MLE, comparing SERA motif normalized enrichment scores using zero-inflated negative binomial (ZINB) or ZI Poisson regression with a log link transformation. Group inclusion is as in (a,b). SERA threshold of positivity is >=3 for individual linear peptides as denoted by dotted line. Values below threshold were given a score of zero. Significance was derived from the incidence rate ratio estimate of each factor in the model. (f) Comparison of select cytokine concentrations amongst sex-based groups. Comparisons were done using Kruskal-Wallis test and significance was adjusted using the Steel-Dwass method (IL-8 and TGF-β2) or ANOVA (APRIL) with Tukey adjustment, as determined by normality of data distribution. (g) Comparison of testosterone levels across sex-based groups. Group inclusion is as in (a,b). Comparisons were done using Kruskal-Wallis with Steel-Dwass adjustment. (h) per-unit odds ratios of hormones associated with Long COVID status amongst all females not on sex-hormone therapies. Odds ratios were obtained separately for each hormone using logistic regression with adjustment for age, BMI, vaccination status and hormone conditions. Hormones with a significant predictive ability are labeled with an asterisk (*). (i) Comparison of estradiol levels across sex-based groups. Group inclusion is as in (a,b). Comparisons were done using Kruskal-Wallis with Steel-Dwass adjustment. (j) per-unit odds ratios of hormones associated with Long COVID status amongst all males not on sex-hormone therapies. Odds ratios were obtained separately for each hormone using nominal logistic regression with adjustment for age, BMI, vaccination status and hormone conditions. Hormones with a significant predictive ability are labeled with an asterisk (*).

### Female-specific immunological features in LC

PLS-DA analysis among females was optimized at three components accounting for a sizeable portion of the variance (cumulative pseudo R-squared = 0.98) and revealed significant differences in the female LC immune signature compared to control females (Figure 2a, and Extended Figure 2a) with 125 features above the initial VIP threshold cutoff criteria and 28 features meeting all bootstrapping CI criteria. Females with LC showed enrichment of an exhausted and effector T cell phenotype with higher levels of CD4 and CD8 T cell exhaustion (CD4Tex, CD8Tex), and a wide array of cytokine-secreting CD4 and CD8 T cells both previously seen in our cohort such as IL-4, IL-6, and IL-4/IL-6 double positive CD4 and CD8 T cells^8^, and some not previously identified including IFN-γ secreting CD4 T cells, suggesting an active and ongoing effector process in females with LC. Females with LC also showed an enrichment of double negative B cells as well as HLA-DR+ B cells and certain factors involved in inflammation and complement including IL-8 and C4b, as well as growth hormone (GH) and neural growth factor (NGF). In addition, females with LC also showed higher antibody levels against SARS-CoV-2 S1 protein, and higher antibody reactivity to a previously identified EBV gp42 motif, PVNFNK elevated in individuals with LC^8^, compared to control females. While some other PAIS and chronic or latent antibody reactivity profiles as well as smith antibodies met initial VIP criteria, they did not pass the bootstrap threshold criteria. Finally, some factors were downregulated in females with LC including cortisol, testosterone, and the mucosal-tissue-associated chemokine CCL28 (Figure 2a, and Extended Data Figure 2a).

### Male-specific immunological features in LC

We performed a similar PLS-DA analysis comparing males with LC with control males. Dimensionality reduction analysis was found to be optimized at three components explaining a high portion of the variance (cumulative pseudo R-squared= 0.98) with 125 features above the initial VIP threshold cutoff criteria and 36 features meeting all bootstrapping CI criteria. Males with LC demonstrated significant differences in innate cellular phenotype and cytokine signatures compared to control males (Figure 2a and Extended Figure 2b). Specifically, males with LC demonstrated a broad reduction of myeloid-derived cell lineages including total monocytes, classical monocytes, low-density neutrophils, and dendritic cell populations including DC1 and pDC. Males with LC showed an increase in the proportion of low-density eosinophils, and NK cells compared to controls. Other immune features significantly enriched in males with LC appeared to be APRIL, CCL20, TGF-β1, and TGF-β2. Similar to females with LC, males with LC showed elevation of IL-8, and antibodies against SARS-CoV-2 S1 and also to total Spike protein. Finally, males with LC showed lower levels of estrogen, and higher levels of insulin, NGF, and oxytocin, compared to their control male counterparts (Figure 2a, and Extended Data Figure 2b).

Comparatively, PLS-DA analysis revealed remarkably distinct LC phenotypes in females and males. While certain factors were shared in LC status irrespective of sex, such as lower cortisol, lower DC1 populations, and higher IL-8 levels, consistent with our initial findings^8^, other factors such as higher NK cells, the depletion of the monocyte populations, increased cytokine-secreting T cells, EBV reactivity and TGF-β were predominantly confined to one of the sexes. Indeed, females and males with LC had distinct top-ranking subclass features associated with Long COVID status (Extended Figure 2a, b).

### LC sex-specific phenotypes in sex-integrated analysis

Following the identification of distinct immune signatures in females and males with LC through sex-stratified analyses, we proceeded with a sex-integrated analysis using PLS-DA. Importantly, the sex-integrated PLS-DA model was designed to analyze sex-based LC status, categorizing participants into four groups: females with LC (LCF), males with LC (LCM), control females (CF), and control males (CM). This categorization allowed the model to differentiate not solely based on sex or LC status alone, but on their interplay.

Dimension reduction for the sex-integrated PLS-DA model to compare sex-based LC status was found to be optimized at seven components accounting for a sizeable portion of the variance (cumulative pseudo-R-squared = 0.89) with 156 features meeting initial VIP threshold criteria and 57 features meeting the bootstrapped VIP threshold criteria. Our sex-integrated analysis once again revealed strong sex differences in the immune phenotypes of females and males with LC. These phenotypes were distinguishable from one another using even the first two of the seven components from our optimized sex-integrated model (Figure 2b). Notably, testosterone was ranked as the most important predictor in this model, over cortisol, as a predictor of sex-based LC status, emphasizing testosterone’s role in the interplay of both LC status and sex (Extended Data Figure 3 a). Differences in the bootstrapped distribution of each feature’s relative regression coefficient for each sex-based group in the PLS-DA model again revealed that females with LC demonstrated enrichment of cytokine-secreting T cells and exhausted CD4 T cells as well as antibody reactivity to EBV and CMV even over males with LC (Extended Data Figure 3 a-d). NK cells, APRIL, and TGF-β family members fell in the quadrant for males with LC and were associated with males with LC over any other sex-based group (Extended Data Figure 3 a-e).

Finally, we compared the differences of the top predictors for sex and disease status obtained from our PLS-DA models as well as antibody reactivity differences in additional herpesviruses such as CMV, HSV-1, HSV-2, and HHV-6b across sex-based groups using standard regression techniques and included (Figure 2 c-f; Extended Data Figure 4, 5). Once again, our sex-predominant findings were recapitulated for these cellular, cytokine, endocrine, and humoral phenotypes seen in females and males with LC, even after accounting for age and BMI (Figure 2 c-f, and Extended Figure 4). Similarly, higher reactivity to CMV and EBV linear epitopes continued to be observed in females with LC even after accounting for vaccination status, age, or inferred prior herpesvirus exposure history (Extended Data Figure 5). We also appreciated an enrichment of total HSV-2 antibody reactivity in females with LC compared to other sex-based groups, even after accounting for age (Figure 2e and Extended Data Figure 5). Notably, females with LC were disproportionately inferred to be HSV-2 positive by antibody reactivity enrichment quantification to a linear peptide display library, known as Serum Epitope Repertoire Analysis (SERA) (Extended Data Figure 5). Thus, like symptom profiles, females and males show a distinct immune signature associated with LC status.

### Distinct herpesvirus reactivity IgG profiles amongst females and males with LC

To further understand the global pattern of antibody reactivity against herpesvirus in our cohort we used hierarchical clustering on linear epitopes identified by SERA antibody reactivity profiling against herpesviruses including EBV, CMV, HSV-1 and HSV-2. Expectedly, clustering analysis revealed four major clusters amongst the linear peptides, separating largely by herpesvirus type, with EBV, HSV-1, HSV-2 and CMV generating their own clusters (Extended Data Figure 6), suggesting a significant ability for these linear peptides to differentiate between each herpesvirus. In addition, reactivity profiles amongst our cohort lead to three major clusters: Cluster 1 which showed enrichment of all four herpesvirus clusters with the highest enrichment of CMV linear epitopes and lower enrichment of HSV-1 linear epitopes; Cluster 2 which showed high reactivity to only EBV-specific epitopes; and Cluster 3 which showed high reactivity to HSV-1 linear epitopes, and lower reactivity to CMV epitopes (Extended Data Figure 6a). Notably, there was a significant difference in the composition of the three clusters by sex-based LC status (Extended Data Figure 6b, p=0.0243). Females with LC were disproportionately enriched amongst clusters 1 and 2 over cluster 3 and showed the highest proportional composition in cluster 1 compared to other groups. Males with LC showed higher proportional enrichment in cluster 3 compared to any other group while female and male controls showed a disproportionate enrichment in cluster 3 (p= 0.0443, Extended Data Figure 6c). These findings suggested that herpesvirus reactivity was distinct across sexes with females with LC showing higher reactivity profiles for EBV, CMV, and HSV-2 linear epitopes simultaneously, while males with LC tended to have a more EBV-enriched reactivity profile.

### Sex-hormone dysregulation in females and males with LC

We sought to confirm the initial findings from our VIP plots identifying testosterone in females and estradiol in males as significantly lower in LC than controls by first analyzing our data within our cohort with the exclusion of the individuals on hormone therapy, or with hormone-altering conditions, using a standard regression approach across all sex-based groups. Analysis revealed that indeed, females with LC had significantly lower testosterone levels than any other group including control females (Figure 2g). To confirm testosterone and other hormones’ ability to distinguish individuals with LC in a sex-stratified manner, we constructed a logistic regression model amongst those not on sex-hormone therapies and adjusted for age, BMI, and other possible confounders in the model, obtaining the odds ratio for each hormone separately (Figure 2h). Notably, only testosterone and cortisol levels in females with LC were significant negative predictors of LC status with testosterone ranking as having a higher negative per unit odds ratio, demonstrating that testosterone remained a top predictor of LC status in females (Figure 2h). Growth hormone (GH) and brain-derived neurotrophic factor (BDNF), however, were significant positive predictors of disease status in females with LC. Similar findings in both linear regression and logistic regression were observed for estradiol in males with LC compared to control males, where estradiol showed a strong negative association with disease status (Figure 2i, j). These results suggested that individuals with LC suffered from a relative deficiency in non-dominant sex hormones and combined with the lower cortisol levels with normal ACTH^8^, suggesting a possible dysregulation of the hypothalamic-pituitary-gonadal (HPG) axis.

### Hormone-independent sex-derived immune signatures can distinguish females and males with LC

Our PLS-DA models suggested that females and males with LC have distinct immune-endocrine signatures. However, females and males naturally vary in their hormone profiles. Therefore, we wondered if the cellular and cytokine signatures alone, devoid of hormone levels, were sufficient to significantly distinguish females from males with LC within our cohort.

To construct these sex-specific immune scores, we extracted 30% of our cohort (n=50) through a random balanced selection of sex and potential confounders and designated this subset as our testing subset, which would be used to test for reproducibility of our models in extracting a distinct sex-based immune signature (Figure 3a). We constructed prediction models for LC status in a sex-stratified manner using PLS-DA and the remaining 70% (n=115) of our cohort, utilizing cellular, cytokines and non-SARS-CoV-2 antibody profiles but devoid of all hormones. A five-fold cross-validation on this training cohort produced a female-derived LC model with five PLS components and a male-derived LC model with three components, achieving cumulative pseudo R-squared values of 0.99 and 0.98, respectively (Figure 3a).

**Figure 3:**
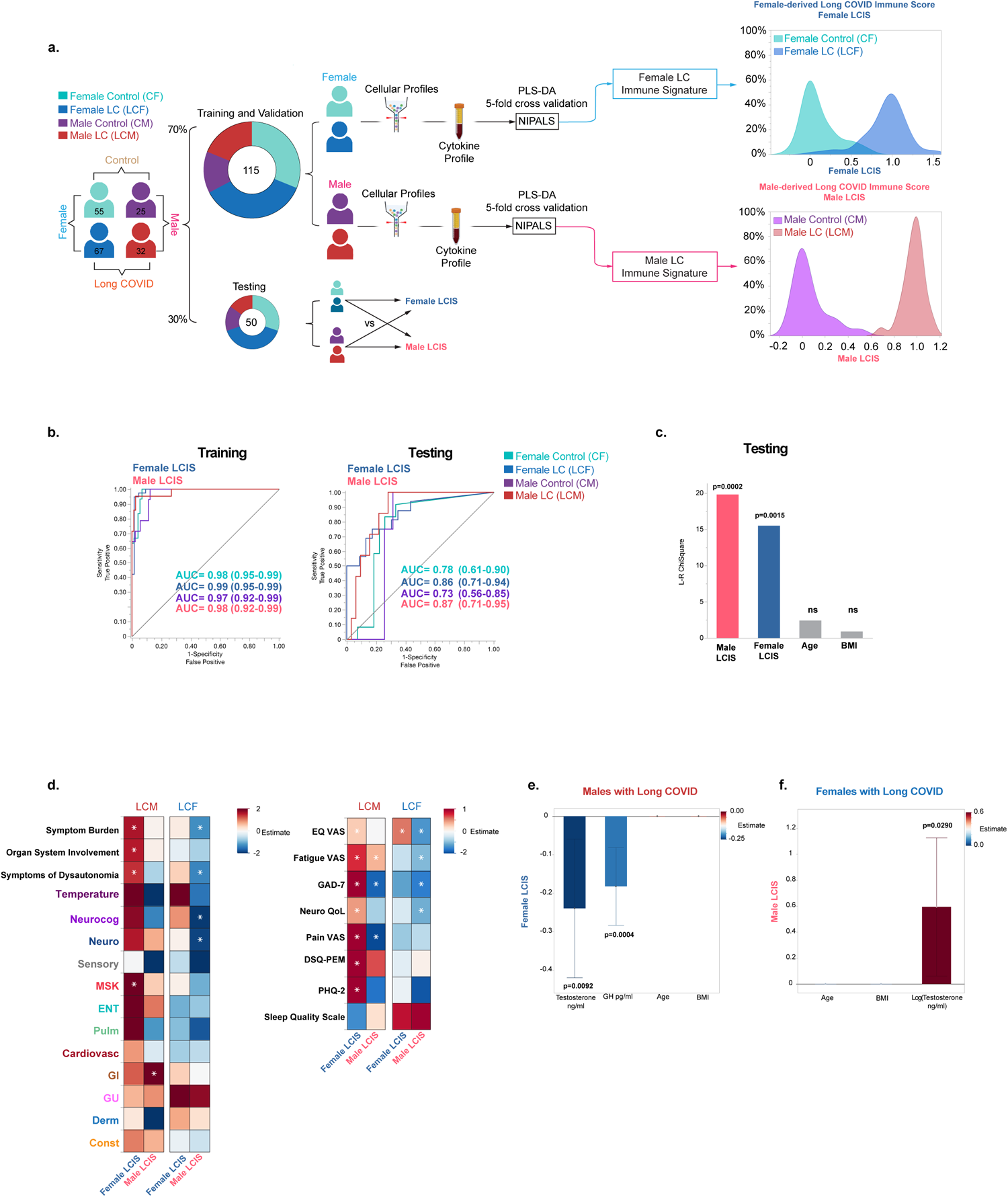
Hormone-independent sex-derived LC immune signatures. a. Schematic depicting the generation of hormone-independent sex-derived immune signatures, a female-derived Long COVID immune score (female LCIS) and a male-derived Long COVID immune score (Male LCIS). Study data was separated into a training and validation subset (70%, n=115) for model building and a testing subset (30%, n=50) through random balanced selection accounting for sex-based group designation, vaccination dose, presence or absence of respiratory conditions, hormone disorders, and hormone or immune modifying therapies. Only the training dataset was used for model building. Feature inclusion into each model was as in Figure 2, except for the exclusion of hormones and antibody responses to SARS-CoV-2. Models were generated using PLS-DA with 5-fold cross validation, for prediction of disease status in a sex-stratified manner. The Female-derived model was optimized at five principal components with a cumulative R-squared =0.99. The male-derived model was optimized at three principal components with a cumulative R-squared =0.98. Prediction Formula for each model was then applied to all member of our study, resulting in a female-derived Long COVID immune score (female LCIS) and a male-derived Long COVID immune score (male LCIS) for each member of our study (females and males; training and testing). Right Panels show the distribution of the female-derived and male-derived immune scores in the full dataset (training and testing) of females and males respectively. b. Receiver-Operator Curve (ROC) analysis of a multinomial Logistic regression model with inclusion of age, BMI, female LCIS, and male LCIS, as predictors of sex-based LC status for training (left) and testing (middle) subsets. Area under the Curve (AUC) for each sex-based group is shown for both the training and testing data with DeLong 95% CIs shown. Right panel shows Likelihood Ratio Chi-Squared Statistic for predictors in the testing model with significant p-values shown. c. Heatmap showing the association of female LCIS and male LCIS on symptoms (left) and standardized health surveys (right panel) in females and males with Long COVID. Each symptom and survey was done separately using GLM with Poisson maximum likelihood estimation and a log link function. Age and BMI were adjusted for in each model. Data is amongst vaccinated individuals without pre-existing respiratory conditions. Asterisks denote significance of the respective LCIS as a predictor of the respective symptom or health survey. (d., e.) Double LASSO with k-fold cross validation (k=5) showing the significant feature predictors of the female LCIS in males with Long COVID (f) and of male LCIS in females with Long COVID (g.) Each model was run using all thirty-two hormones, peptides, and myokines not used in the generation of the Long COVID immune scores. Figures show only non-zero factors along with age and BMI.

We then applied the scoring formula for each sex-derived model separately to all members of our study, which could be used to measure the relative presence of the immune signatures obtained from females and males with LC in each individual (a female-derived Long COVID Immune Score [Female-LCIS]; and male-derived Long COVID Immune Score [Male-LCIS], Figure 3a). Receiver-Operator Curve (ROC) analysis revealed a significant predictive ability of the Female-LCIS to distinguish between females with LC and control females for both the training and the testing subsets of our cohort (AUC= 1.0, training; AUC = 0.88, 0.70-0.96 95% CI) testing; p< 0.0001 Figure 3b). Similarly, the Male-LCIS showed a strong ability to predict LC status in males in both the training and testing subsets of our cohort (AUC= 1.0, training; AUC =1.0, testing, p< 0.0001, Figure 3c). Finally, we ran a multinomial logistic regression model using the female and male-derived LC immune scores on both females and males in a sex-integrated manner, to understand if these two immune scores could distinguish between the four sex-based groups. ROC analysis demonstrated that these two independently derived immune scores were sufficient to significantly distinguish females with LC, males with LC, control females, and control males from each other in either our training or testing groups when all individuals were included and even when accounting for age and BMI (Figure 3d). These results demonstrated that the immune signatures found in females and males with LC were significantly different and could reproducibly predict both the sex and disease status of individuals, even without knowing their hormone levels.

### Hormone-independent sex-derived LC immune signatures predict symptom burden

Given our findings, we wondered if sex-specific immune signatures were associated with the distinct symptom profiles we uncovered in females and males with LC. To analyze this, we ran Poisson regression on our cohort in a sex-stratified manner, among vaccinated individuals not on sex hormone therapy and without pre-existing respiratory conditions. We compared the relationship between the two sex-derived LC immune scores (Female LCIS and Male LCIS) and various symptom dimensions including symptom burden, organ system involvement, the total number of symptoms within each organ system, and responses to the standardized quality of life surveys while accounting for age and BMI. The results revealed that sex-derived LC immune signatures were indeed correlated to symptom patterns for both sexes (Figure 3e). Males with LC showed significant associations to poorer health quality and higher symptom burden with a higher female-derived LC immune score. For example, males with LC reported significantly higher symptom burden, organ system involvement, and showed higher neurocognitive impact on quality of life, and greater fatigue with an increasing female-derived LC immune score, suggesting that the increased prevalence and reporting of many of these symptoms in females with LC was indeed reflected in their immune signature. Similarly, females with LC who exhibited a higher male-derived LC immune signature demonstrated significantly lower symptom burden, neurocognitive, and neurological symptoms and reported lower neurological impact on quality of life, and lower fatigue (Figure 3e). These findings suggest that the sex-specific differences in the immune response to LC in females and males are dynamically associated with their LC symptom profiles in both sexes.

### Testosterone is a key correlate of the sex-specific immune phenotypes

Given our initial findings that the non-dominant sex hormones of females and males with LC were significantly lower compared to their control counterparts, we sought to understand if hormones themselves may help explain the distinct immune phenotype that we observed in females and males. To test this relationship, we took advantage of the sex-derived LC immune signatures, which were derived independent of hormones, allowing us to test their relationship in a non-circular manner. Regression analysis revealed testosterone as the top predictor of the male-derived LC immune score in females with LC, which was in turn associated with lower symptom profiles in females with LC (Figure 3f). Notably Female-LCIS in males with LC, positively associated with higher symptomology in males with LC, was negatively predicted by testosterone levels as the top negative predictor followed by growth hormone, suggesting once again the involvement of the HPG axis in the immune phenotype of LC across sexes (Figure 3 g). Taken together these data support the concept that the immune signatures in females and males with LC, which were dynamically associated with their symptom profiles are linked to their hormone expression profiles, with testosterone as the top predictor.

### Testosterone levels correlate with male-like immune phenotypes in females

Building on our findings regarding testosterone and its association with the distinct female and male LC immune signatures, we sought to gain granularity in its relationship with each cellular and cytokine feature. To do so, we applied PLS analysis in a sex-stratified manner to find the top predictors of lower or higher testosterone levels in females and males with LC. Testosterone expression patterns in females with LC showed a striking resemblance to the differential expression of the sex-specific immune signatures seen in females and males with LC (Figure 4a). Many of the factors associated with relatively higher testosterone levels in females with LC were those that were differentially upregulated in males with LC relative to control males including TGF-β1, TGF-β2, April, CCL3, and IL-8. Predictors of lower testosterone in females with LC included higher levels of cytokine-secreting T cells such as IL-4 IL-6 double-positive T cells with IL-4-secreting CD4 T cells passing all bootstrapped threshold criteria (Figure 4a and Extended Figure 7a). Lower testosterone levels in females with LC were also associated with higher reactivity to herpes virus epitopes belonging to EBV and CMV with CMV epitope to UL146 viral CXCL1 (vCXCL1) meeting all threshold criteria.

**Figure 4:**
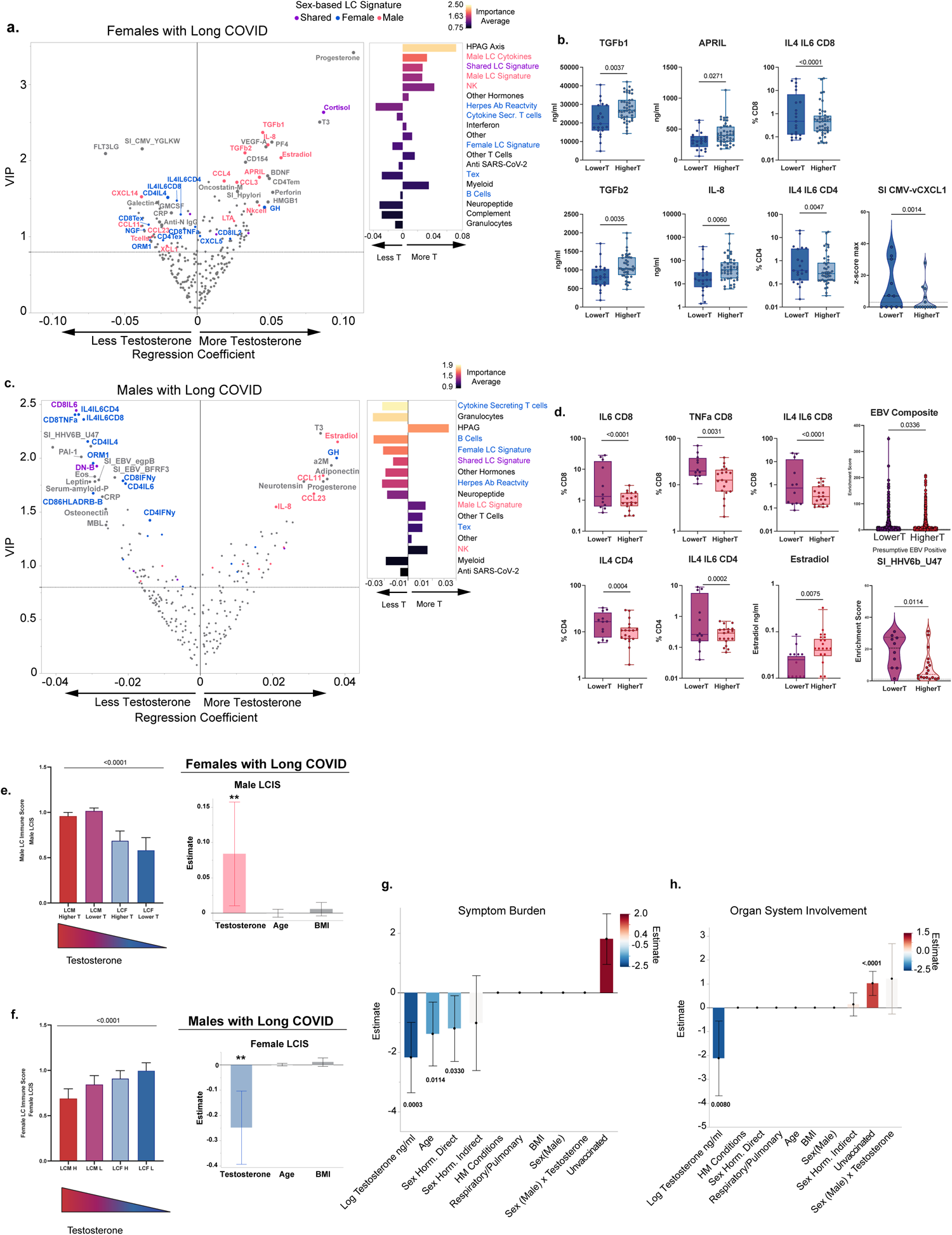
Testosterone levels correlate with male-like LC immune phenotypes in both sexes. (a,c). Sex-stratified Variable importance Projection (VIP) by regression plots for prediction of testosterone levels amongst females with LC (a) and males with LC (b), without pre-existing autoimmune or hormone conditions or sex-hormone therapy. Graphs show VIP Plots (Left) and normalized summative regression scores by grouping classification (right). Normalized summative regression scores are color-coded by the Importance Average of the group, defined as the VIP score average for all members of the group. VIP by regression Plots were derived from PLS analysis with 5-fold cross validation. VIP and regression scores for each feature were bootstrapped to generate 95% CIs. Dotted line denotes the threshold cutoff (0.8) for feature importance. Regression is denoted on the x-axis with a solid line indicating the intercept. Positive values indicate positive predictors of testosterone and negative values indicate negative predictors of testosterone. Each dot represents a unique feature included in the model. Color-coding denotes sex predominance (as previously determined in Figure 2) for Long COVID status for all features passing the importance threshold. Labeling is shown for all features passing all bootstrap threshold criteria (larger circles), and sex-predominant Long COVID features passing initial threshold criteria. Additional details of the PLS analyses can be found in Extended Figures 7 and 8. (b, d) Comparison of select features amongst the bottom tercile of testosterone levels (Lower T) and the top two terciles of testosterone levels (Higher T) in females with LC (b), or males with LC (d). Cytokine and hormone expression levels were done using standard t-test or Mann-Whitney test as appropriate. Cell frequencies were analyzed using Poisson regression or negative binomial regression. SERA analysis was done using zero-inflated (ZI) Poisson or ZI negative binomial regression. Benjamini FDR-adjusted p-values are shown. EBV composite score (d, top right) is the composite of all EBV epitopes measured by SERA for which expression was available amongst both groups. Analysis was done using generalized linear mixed models (GLMM) with Poisson distribution accounting for each individual and each epitope as a repeated measure. (e) Comparison of male-derived (top) and female-derived (bottom) Long COVID immune scores amongst Sex and testosterone expression groups. Groups were ordered from left to right based on decreasing testosterone expression. Left panels show the linear trend of the mean across the groups. Right panels show the generalized linear regression models comparing Testosterone levels and Male LC Immune Scores (Male LCIS) amongst females with LC (top) or testosterone levels and Female LC Immune Scores (Female LCIS) amongst males with LC (bottom) without pre-existing hormone conditions or sex-hormone therapy, adjusted for age and BMI. (h,i) adaptive double LASSO with Poisson distribution and loglink function identifying predictors of symptom burden (h) and organ system involvement (i) in a sex-integrated manner comparing testosterone levels, sex designation, their interaction, and other confounders as listed.

To confirm the immune factors found through PLS-DA modeling were indeed associated with testosterone levels in females with LC, we divided females with LC into two groups: a relatively lower testosterone group (Lower T, n=24) representing the bottom tercile of testosterone expression amongst females, and a relatively higher testosterone group (Higher T, n=33) representing the top two terciles of testosterone expression amongst females. This stratification was chosen to better reflect the distribution of testosterone expression amongst our female cohort and provide a consistent criterion that could in turn be applied to males with LC (Extended Data Figure 8a). We compared the expression levels of the top male predictors TGF-β1, TGF-β2, APRIL, and IL-8 among these two groups and found that indeed Higher T females with LC had significantly higher expression levels of these factors that were differentially expressed in males with LC. In addition, females with LC at the bottom tercile of testosterone expression showed higher linear epitope reactivity to CMV-UL146 (vCXCL1); and even higher expression of IL-4/IL-6 double-positive CD4 and CD8 T cells (Figure 4b). Similar findings were observed when accounting for age, BMI, and other confounders, or when an alternate division point was chosen for females and males with LC separately (Extended Data Figure 8 b-e). Together these findings suggested that testosterone levels in females with LC were differentially associated with the spectrum of immune factors found in either females or males with LC, with higher testosterone levels showing higher activation of factors found in males with LC, including TGF-β and APRIL, and lower testosterone associated with factors found in females with LC including higher levels of cytokine secreting T cells and increased antibody reactivity to herpesviruses such as EBV and CMV.

### Testosterone levels correlate with male-like immune phenotypes in males

The findings in females with LC prompted us to do the same in males with LC, predicting testosterone levels using their immune factors (Figure 4c and Extended Data Figure 7b). Strikingly, predictors of lower testosterone in males with LC were factors that were predominant in the immune signature of females with LC. Lower testosterone levels in males with LC were predicted by higher cytokine-secreting CD4 and CD8 T cells including IL4, IL6, TNF-α, and IFN-γ as well as the IL-4/IL-6 double-positive CD4 and CD8 T cell populations. Higher antibody reactivity to several herpesvirus epitopes, particularly to EBV and HHV6b were also predictive of lower testosterone levels in males with LC. By contrast, higher levels of NK cells, CCL23, ADAMTS13, and IL-8 were predictive of higher testosterone levels in males with LC, with IL-8 passing all bootstrapping criteria. Finally consistent with the pattern observed in females with LC, higher testosterone levels in males with LC were associated with increased levels of HPG hormone signaling, including higher levels of growth hormone, estradiol, progesterone, and cortisol, with GH and the sex hormones passing all bootstrap threshold criteria (Figure 4c and Extended Data Figure 7b).

Like with females, we separated males into relative expression groups of testosterone with the bottom tercile expression labeled as Lower T (n=12), and the top two terciles labeled as higher T (n=19) (Extended Data Figure 8a). We then compared some of the top features identified by PLS-DA as associated with testosterone levels (Figure 4d). Consistently, males with LC in the Lower T group showed significantly higher expression levels of cytokine-secreting T cells including IL-6 and TNF-α-secreting CD8 T cells as well as IL-4/IL-6 double positive CD4 and CD8 T cells and IL-4 secreting CD4 T cells. Lower T males with LC also showed significantly higher antibody reactivity to a linear epitope to HHV6-b-U47. Similar findings were observed when accounting for age, BMI and other confounders, or alternative cutoffs (Extended Data Figure 8d, e). In addition, Lower T males with LC showed significantly higher average enrichment of antibodies against total EBV linear epitopes (Figure 4 d and Extended Data Figure 8f). Higher T males with LC showed significantly higher levels of estradiol (Figure 4d). Once again, these findings in males with LC suggested that testosterone levels showed a key relationship in the sex-based immune signature found in both females and males with LC.

With testosterone as a key differentiator, we again compared the four testosterone-sex long COVID groups (Male: Higher T, Lower T; Female: Higher T, Lower T) for their relative expression of female-derived and male-derived LC immune scores. Indeed, the female and male LCIS demonstrated a significant positive and negative trend to the declining testosterone level in each of these four groups respectively (Figure 4 e, f). Further in females with LC, higher testosterone levels were positively associated with a higher male-derived LC immune score while in males with LC, lower testosterone was associated with higher scoring female-derived LC immune scores. This trend persisted with key features identified by PLS-DA including subsets of cytokine-secreting T cells and TGF-β signaling (Extended Data Figure 8 g-i). For herpesvirus antibody reactivity, however, clear sex-based differences existed in being associated with testosterone levels. While antibody reactivity to EBV and HSV-2 linear peptides showed a negative correlation with testosterone in both sexes, antibody reactivity to CMV was significantly associated with lower testosterone levels only in females with LC, while antibody reactivity to HHV6b and HSV-1 showed this effect in males with LC (Extended Figure 8j).

### Testosterone levels predict symptom burden in LC over sex designation

The profound immuno-endocrine differences in females and males with LC, with testosterone as a key correlate, prompted us to ask if testosterone levels may be related to the distinctions in symptomology across sexes. To understand this, we tested the association of testosterone levels with symptom burden and organ system involvement in a sex-integrated manner while accounting for sex, testosterone levels, the interaction of sex with testosterone, BMI, and age as well as other possible confounders including vaccination status, as predictors of symptom burden and organ system involvement. Unvaccinated status remained a positive predictor of symptom burden (Figure 4g). Notably testosterone levels could significantly predict symptom burden and organ system involvement in individuals with LC irrespective of sex classification (Figure 4g, h). That is, after accounting for testosterone levels, sex designation was no longer a significant predictor of symptom burden or organ system involvement in individuals with LC. These findings connect the immuno-endocrine phenotype observed in LC to symptomology in LC, most specifically to the symptom burden and organ system involvement.

## Discussion

Our study sought to explore the interplay between symptomology and immunoendocrine function in LC by considering sex as a biological variable. Using participant survey data and a wide immunoendocrine profile, we leveraged machine learning to understand if and how the experience of LC across sexes may manifest biologically. Our approach has identified that both the experience of LC and its immunological manifestations are vastly different between sexes. Females with LC experience a higher symptom burden than males and have a distinct, reproducible, and extractable immune endocrine profile that is reflective of their symptom manifestations. Specifically, the LC immune signature obtained from females, when partially reflected in males with LC was associated with the reporting of higher symptom burden, higher neurological impact as well as higher reporting of pain, fatigue, changes to mood, higher anxiety, and higher post exertional malaise in these males. Conversely, females who partially reflected the immune signature seen in males with LC showed lower symptom burden including lower neurological and neurocognitive symptoms, less fatigue and less anxiety, suggesting that the higher reporting of symptoms in females with LC can at least in part be attributed to the immune-endocrine signature that they exhibit. In addition, we have identified testosterone as a key player in many of these sex differences such as in herpesvirus reactivity.

In our recent study on LC with sex-aggregated analyses^8^, we observed a significant increase in the antibody levels against EBV, suggesting the possibility of reactivation of this latent virus in individuals with LC. Strikingly, in this sex-stratified study, we uncovered that the elevation of the antibody levels against EBV, CMV, and HSV-2 is predominantly observed in females among those with LC. EBV infection is associated with the pathogenesis of autoimmune diseases such as multiple sclerosis (MS) and systemic lupus erythematosus (SLE)^19–22^. Interestingly, levels of androgens such as DHT, DHEA, and DHEA-S, are downregulated in females with SLE compared to age-matched control females^23–25^. In our study, lower testosterone levels in females were associated with higher antibody reactivity to EBV, and CMV, while in males lower testosterone was associated with higher reactivity to EBV and HHV6-b, suggesting that testosterone levels may play a crucial role in maintaining the latency of herpesviruses and in turn to the predisposition to autoimmune conditions, such as SLE and MS.

Previous studies have shown that androgens, including testosterone, have immunosuppressive roles, which could be potentially associated with the strong female bias in autoimmune diseases^26^. Androgen receptors are expressed in a wide range of immune cells, from myeloid cells such as macrophages/monocytes, neutrophils, mast cells, eosinophils, to lymphoid cells including B cells and T cells^26–29^. This vast range of cell types that can be modulated by testosterone therefore suggests that testosterone can lead to profound changes in the immune landscape of an individual. We found that females and males with LC had a relative decrease in their non-dominant sex hormones. We observed a significant and profound association in the immune phenotype of females and males with LC that was strongly tied to the levels of testosterone in both sexes. Lower testosterone levels in either sex were strongly associated with the immune phenotype predominantly observed in females with LC which included a strong T cell-mediated response. It has been shown that androgen deprivation leads to elevated T cell levels and enhanced responses^30^. By contrast, higher levels of testosterone were associated with a lower T cell response and higher signaling of factors such as APRIL and TGF-β family members and NK cells which have been previously shown to be associated with testosterone^31–33^. Notably, in adult human males, estradiol is generated in large part (approximately 80%) from the aromatization of testosterone through the enzyme aromatase, found in several key tissues including the gonads, brain, and adipose tissues. It is therefore possible that our observation of lower circulating estradiol levels in males with LC may be a consequence of lower aromatase activities in key tissues. Future work in studying these hormones in females and males may help shed light on this concept.

The deep immune-endocrine profiling in our study has provided us the opportunity to understand the breadth of association that this testosterone has in the immune system. In addition, this hormone was also associated with the distinct symptom phenotypes observed across sexes and testosterone was able to override sex designation as a predictor of symptom burden. These findings provide an avenue for possible therapy to mitigate the severity of LC symptoms through hormonal replacement therapy. Interestingly hormone therapies were among the factors predicted as negative contributors of symptom burden in our study.

Collectively, our findings suggest that individuals with LC may suffer from a dysregulation of key neuro-endocrine and other endocrine disorders, which are in turn associated with herpesvirus reactivity. It should be noted that LC symptoms have a significant overlap with those found in other PAIS conditions^5^. These findings provide support to investigate the role of sex hormones in PAIS including ME/CFS and chronic Lyme disease^11,34–37^.

In addition to previously described limitations^8^, another limitation of this work is in its compositional diversity in race and ethnicity as well as in the number of individuals with greater variability in gender identity that may be distinct from their assigned sex at birth. For example, in addition to cis-gendered females, trans-gendered individuals show an even higher rate of LC^42^. Whether this higher rate is due to hormonal therapies for those who elect to undergo physical transitional changes that better match their gender identity (e.g. inhibition of testosterone among some trans individuals), or due to systemic societal issues remains unknown. Greater granularity in understanding this population will likely lead to better insights into our understanding of biological sex differences, social disparities, and the interplay of these two facets of the immune landscape.

Our work suggests that dysregulated control of hormones downstream of the hypothalamus-pituitary-organ axis may be a common feature of individuals with LC. Our work suggests that sex differences in LC, as they pertain to the immune system, is not an unmovable dichotomy but a dynamic spectrum of which hormones like testosterone may help dictate the immune landscape and its approach to a perceived threat. These findings inform diagnostic and therapeutic strategies that may correct or alleviate the neuroendocrine deviation caused by the immune system.

## Methods

### Ethics statement

This study was approved by the Mount Sinai Program for the Protection of Human Subjects (IRB #20-01758) and Yale Institutional Review Board (IRB #2000029451 for MY-LC; IRB #2000028924 for enrollment of pre-vaccinated Healthy Controls; HIC #2000026109 for External LC). Informed consent was obtained from all enrolled participants.

### MY-LC Study Design, Enrollment Strategy, and Inclusion / Exclusion Criteria

The MY-LC study was a multi-center, cross-sectional research project as previously described. It encompassed five distinct groups, each with different histories of exposure to SARS-CoV-2 and varying manifestations of Long COVID, as described previously^8^. For inclusion in the Long COVID cohort, candidates underwent thorough medical evaluations to exclude other possible causes for their lingering symptoms.

Recruitment channels for participants with ongoing symptoms post-COVID-19 included the Long COVID clinics within Mount Sinai’s network and the Mount Sinai Hospital’s Center for Post COVID Care. Those in the healthy and convalescent groups were reached through IRB-sanctioned promotional methods, including emails, flyers in hospital areas, and social media outreach. Every participant gave informed consent at enrollment. Data collection involved gathering peripheral blood samples and symptom survey responses on the day of sampling, alongside a review of self-reported medical histories, supplemented by electronic medical record analysis by our team of clinicians.

#### Inclusion and Exclusion Criteria

Eligibility for the Long COVID group (LC) required being 18 years or older, a past confirmed or probable COVID-19 infection as per WHO guidelines, and experiencing symptoms for over six weeks post-infection. Healthy control (HC) candidates were 18 or older, with no history of COVID-19, confirmed through a brief screening. The convalescent control group (CC) had similar age and past infection criteria, with an additional screening confirming the absence of current symptoms.

Exclusion criteria encompassed inability to consent and conditions precluding blood tests. Post-enrollment, participants could be excluded based on pregnancy, significant immunosuppression (exceeding prednisone 5 mg daily), active cancer or chemotherapy, or specific genetic disorders that could confound results. In addition, for certain immunological assessments, participants with conditions or therapies that could confound results (like sex hormone therapy for measures of sex hormones) were also excluded. These specific exclusions are denoted in our figures and detailed in the accompanying text and legends.

### Participant Surveys

In the MY-LC study, an extensive array of surveys was utilized to gather data from participants as detailed prior. This included a combination of established patient-reported outcomes (PROs) and bespoke survey tools specifically crafted by our research team. Initial demographic information obtained through these surveys encompassed details like gender, age, BMI, ethnicity, and existing health conditions. Participants from the Long COVID and convalescent groups additionally provided details on their COVID-19 experience, including symptom onset, severity of the acute phase (distinguishing between hospitalized and non-hospitalized cases), results from SARS-CoV-2 PCR tests, and antibody tests. All study participants were also queried about their vaccination status against SARS-CoV-2, including the dates and types of vaccines received.

On the day of blood sample collection, participants completed various PROs to assess different health aspects. These assessments included the Fatigue Severity Scale (FSS)3 and a fatigue visual analogue scale (F-VAS) for fatigue, the DePaul Symptom Questionnaire Post-Exertional Malaise Short Form (DSQ-PEM Short Form)4 for post-exertional malaise, the Medical Research Council (MRC) Breathlessness Scale5 for breathlessness, the Neuro-QOL v2.0 Cognitive Function Short Form6 for cognitive health, the EuroQol EQ-5D-5L7 for health-related quality of life, the GAD-78 for anxiety, the PHQ-29 for depression, a pain visual analogue scale (P-VAS), and the Single-Item Sleep Quality Scale10 for sleep quality. Additionally, information regarding employment status before and after COVID-19 was collected using a survey developed by our authors. Participants were also invited to list any ongoing symptoms from a provided standardized list.

### Calculation of Symptom Burden and Organ System Involvement

#### Symptom Burden

Symptom burden in this study is defined as to total number of ongoing symptoms experienced by a given individual from a standardized list of x symptom options provided to each participant at the time of sample collection as described above. Symptom burden was calculated in two ways: First, for a sex-independent calculation of symptom burden that could be used to compare across males and females with LC, each individual’s total symptoms -with the exception of sex-specific symptoms such as changes in menstrual cycle--were summed. For a sex-stratified calculation of symptom burden, all symptoms including those that are sex-specific were counted for each individual and summed. This calculation of symptom burden was used for within-sex comparisons.

#### Organ System Involvement

Each of the 42 symptoms on the standardized symptoms questionnaire was classified into one of 12 organ systems per the mutual consensus of at least two practicing physicians (see Extended Data Table 3). Organ system involvement was defined as the sum of distinct organ systems reported by an individual per the classification of each of their ongoing symptoms.

For secure and efficient data collection and storage, all survey responses were managed using the REDCap (Research Electronic Data Capture) tools, hosted by the Mount Sinai Health System.

### Blood Sample Processing

Blood sample processing was done as detailed before. Briefly, whole blood from participants was collected in sodium-heparin-coated vacutainers (BD 367874, BD Biosciences) at Mount Sinai Hospital in New York City, New York. All participant samples were assigned unique MY-LC study identifiers and de-identified by clinical staff. On the same day of sample collection, samples were transported directly to Yale University in New Haven, CT and processed on the same day. Plasma samples were collected after centrifugation of whole blood at 600×g for 10 minutes at room temperature (RT) without brake. Plasma was then transferred to 15-mL polypropylene conical tubes, aliquoted, and stored at −80°C. The peripheral blood mononuclear cell (PBMC) layer was isolated using SepMate tubes (StemCell) according to the manufacturer’s instructions. Cells were washed with phosphate-buffered saline (PBS), pelleted, and briefly treated with ACK lysis buffer (ThermoFisher) for 2 minutes before counting. Viability was estimated using standard Trypan blue staining and a Countess II automated cell counter (ThermoFisher). PBMCs were plated directly for flow cytometry studies or stored at −80°C in freezing media. Plasma samples from the External Long COVID group were obtained using BD Vacutainer CPT tubes (#362753) according to manufacturer’s instructions and stored in aliquots at −80°C prior to analysis.

### Flow cytometry

Flow cytometry protocol was done as before. Freshly isolated PBMCs were allocated at a density of 1–2 × 10^6^ cells per well into 96-well U-bottom plates for analysis. The cells were initially treated with Live/Dead Fixable Aqua dye (ThermoFisher) for a 20-minute duration at 4°C. Post-staining, a PBS wash was performed, followed by a 10-minute incubation at room temperature with Human TruStain FcX (BioLegend). Subsequently, we added a mixture of specific staining antibodies to the cells and allowed this to incubate for 30 minutes at room temperature. Before proceeding to the flow cytometry analysis, cells were washed again and then fixed in 100 μl of 4% paraformaldehyde for 30 minutes at 4°C.

For the assessment of intracellular cytokines, cells that had been stained for surface markers were suspended in 200 μl of complete RPMI medium (cRPMI, containing RPMI-1640 with 10% FBS, 2 mM L-glutamine, 100 U/ml penicillin, 100 mg/ml streptomycin, and 1 mM sodium pyruvate) and kept at 4°C overnight. The following day, these cells were further stimulated with 1× Cell Stimulation Cocktail (eBioscience) in 200 μl cRPMI for one hour at 37°C. This was followed by an additional four-hour incubation period at 37°C with 50 μl of 5× Stimulation Cocktail in cRPMI (including a protein transport inhibitor from eBioscience). After this stimulation phase, the cells were washed and fixed again in 100 μl of 4% paraformaldehyde for 30 minutes at 4°C.

To proceed with intracellular cytokine quantification, the cells were permeabilized using the 1× permeabilization buffer from the FOXP3/Transcription Factor Staining Buffer Set (eBioscience) for 10 minutes at 4°C. Staining cocktails for subsequent steps were prepared using this buffer. After permeabilization, cells were washed and incubated with a mixture containing Human TruStain FcX (BioLegend) for 10 minutes at 4°C. Intracellular staining cocktails were then applied to the cells for one hour at 4°C. Following this staining period, the cells underwent a final wash and were readied for flow cytometric analysis using the Attune NXT system (ThermoFisher). Data from this process were evaluated using FlowJo software version 10.8 (BD). Specific antibodies used in this procedure are detailed in Supplemental Table x.

### Multiplex proteomic analysis

Frozen participant plasma was shipped to Eve Technologies (Calgary, Alberta, Canada) on dry ice to run 13 multiplex panels: Human Cytokine/Chemokine 71-plex Discovery Assay (HD71), Human Cytokine P3 Assay (HCYP3-07), Human Cytokine Panel 4 Assay (HCYP4-19), Human Complement Panel Assay (HDCMP1), Human Myokine Assay (HMYOMAG-10), Human Neuropeptide Assay (HNPMAG-05), Human Pituitary Assay (HPTP1), Human Adipokine Panel 2 Assay (HADK2-03), Human Cardiovascular Disease Panel Assay (HDCVD9), Human CVD2 Assay (HCVD2-8), Steroid/Thyroid 6plex Discovery Assay (STTHD) Human Adipokine Assay (HDADK5), and TGF-β3-plex Discovery Assay (TGF-β1-3). Samples were sent in two batches with internal controls in each shipment to assess effectiveness of batch correction as described below. To harmonize data across the two batches, ComBat was used, an empirical Bayes method available through the “sva” R package (version 3.4.6), designating the initial batch as the reference and incorporating the following covariates: disease status, sex, age, and hormone conditions. The effectiveness of ComBat was validated using sample replicates between each batch in a matched pairs analysis. Analytes that exhibited significant differences post-correction were excluded from further analysis.

#### Linear Peptide Profiling

##### SERA plasma screening and inferred herpesvirus positivity

A detailed description of the SERA assay has been published, and has been previously described^38^. For this study, plasma was incubated with a fully random 12-mer bacterial display peptide library (1□×□1010 diversity, 10-fold oversampled) at a 1:25 dilution in a 96-well, deep well plate format. Antibody-bound bacterial clones were selected with 50□µ□ Protein A/G Sera-Mag SpeedBeads (GE Life Sciences, #17152104010350) (IgG). The selected bacterial pools were resuspended in growth media and incubated at 37□°C shaking overnight at 300 RPM to propagate the bacteria. Plasmid purification, PCR amplification of peptide-encoding DNA and barcoding with well-specific indices was performed as described. Samples were normalized to a final concentration of 4 nM for each pool and run on the Illumina NextSeq500. Every 96-well plate of samples processed for this study contained healthy control run standards to assess and evaluate assay reproducibility and possible batch effects. Herpesvirus-specific motifs including to EBV, CMV, and HHV6b, were identified by the IMUNE algorithm as previously described^39^. EBV-specific and CMV-specific motif panels were previously discovered and verified on CMV and EMV predicate positive and negative serum samples respectively. For determining presumptive EBV and CMV positivity in our cohort, participant plasma was screened for enrichment of these pre-identified EBV-specific and CMV-specific motif panels and compared to each panel’s established threshold criteria for positivity. Those meeting the established criteria for the respective panel were classified as presumptive positive.

#### Machine Learning

##### Supervised Dimensionality Reduction Analysis

Partial Least Squares (PLS) and PLS Discriminant Analysis (PLS-DA)

To investigate sex-based differences across symptom and immunoendocrine profiles in our cohort we employed Partial Least Squares (PLS) and PLS Discriminant Analysis (PLS-DA) as supervised data dimensionality reduction techniques. PLS and PLS-DA were used to extract significant features as predictors that explain a continuous variable of interest (PLS model) or discriminate between predefined groups (PLS-DA).

To identify key predictors of testosterone levels, symptomology, and immunoendocrine profiles, analyzed both in sex-stratified and sex-integrated manners PLS and PLS-DA were run using the Non-linear Iterative Partial Least Squares (NIPALS) algorithm with 5-fold cross-validation. PLS and PLS-DA were conducted using JMP® Pro 17.0.0. The final model for each analysis was selected to the optimal number of principal components chosen by the minimization of the Van der Voet’s T-squared statistic and the Root Mean predicted residual error sum of squares (PRESS). Detailed Outputs for each model can be found in Supplementary Table x.

##### Variable Importance on Projection (VIP) by Regression Plots

Post-analysis with PLS and PLS-DA, Variable Importance on Projection (VIP) scores for each feature, which quantify each feature’s contribution to the model’s predictive capacity, were generated and bootstrapped. Features above the VIP threshold cutoff of 0.8, corresponding to the standard threshold for importance,^40^ were considered important. In addition each feature’s regression coefficient, which indicate the direction and magnitude of the relationship between each feature and the response variable, were also generated and bootstrapped. Only features with bootstrapped 95% confidence intervals above the threshold cutoff of 0.8 and with regression 95% CI that did not span the intercept were considered most significant and important. Feature inclusion for each analysis is detailed in accompanying text, figure legends, and supplementary tables. For inclusion of SARS-CoV-2 specific antibodies in the machine learning pipeline, antibody levels were first adjusted by vaccine dose through the inclusion of the residual values after Least Squares mean vaccine dose comparison.

#### General Statistical Analysis

The sample sizes in this study were not pre-determined on a priori power calculations. In general, for the analysis of immunoendocrine plasma-derived features, including systemic cytokine levels, hormone concentrations, and antibody titers, comparisons across subgroups were conducted using mean estimates for normally distributed data or for data that conformed to normality following logarithmic transformation. Classical Analysis of Variance (ANOVA) were applied, with Tukey’s adjustment for multiple comparisons to determine significance. Otherwise, non-parametric methods were utilized. Specifically, the Kruskal-Wallis test was employed as an initial assessment of significant differences across groups, with post-hoc pairwise comparisons using the Steel-Dwass method to adjust for multiple comparisons. For symptom burden, organ system involvement and frequencies of symptoms classified by organ system, and for flow cytometry cell frequency data, groups were compared using generalized linear models (GLMs) with maximum likelihood estimation using Poisson distribution or negative binomial distribution when data was overdispersed. For SERA Enrichment scores for a specific protein, motif, or combined protein group, zero-inflated Poisson or ZI negative binomial regression was used when data was overdispersed. Detailed descriptions of the statistical methodologies employed are provided within the legends accompanying each figure and throughout the text of the manuscript.

## Data availability

All of the raw fcs files for the flow cytometry analysis are available at the FlowRepository platform (http://flowrepository.org/) under Repository ID: FR-FCM-Z6KL.

## Code availability

Computer codes are available as indicated (e.g., https://github.com/xxxx) or otherwise available upon request.

**Additional References (include references used in the Extended Data sections)**

## Acknowledgments

The authors would like to express their immense gratitude to the community of MY-LC study participants who volunteered both their time and effort in aiding the completion of this study, and who also helped inform and educate on the effective and equitable communication of the results from this study. Various graphical schematics were created with BioRender.com. This work was supported by the Else Kröner Fresenius Prize for Medical Research 2023, grants from the National Institute of Allergy and Infectious Diseases (R01AI157488 to A.I.), FDA Office of Women’s Health Research Centers of Excellence in Regulatory Science and Innovation (CERSI) (to A.I.), RTW Foundation (D.P.), the Howard Hughes Medical Institute Collaborative COVID-19 Initiative (to A.I. and R.M.), and the Howard Hughes Medical Institute (to A.I. and R.M.). Steve and Alexandra Cohen Foundation (D.P. and J.W.), Nash Family Foundation (D.P. and J.W.) and Polybio Research Foundation (D.P. and J.W.)

## Author contributions

Experimental conceptualization, methodology, and data visualization were performed by AI, DP, JS. TT, LG; formal analysis conducted by JS, TT; resources provided by D.V.D., D.P., and A.I.; clinical review of electronic health records was performed by JW, JG, and JS.; sample collection, processing, and biospecimen validation were performed by ST, JS, PL, MP; writing – original draft by JS. AI. T.T.; writing – review & editing by JS, AI, TT, JW, DVD, LG, CL, ST, and LG; funding acquisition by AI.

## Additional Information

Correspondence and requests for materials should be addressed to: A.I., L.G. and D.P.

## Extended Data Figure Legends

**Extended Data Figure 1:**
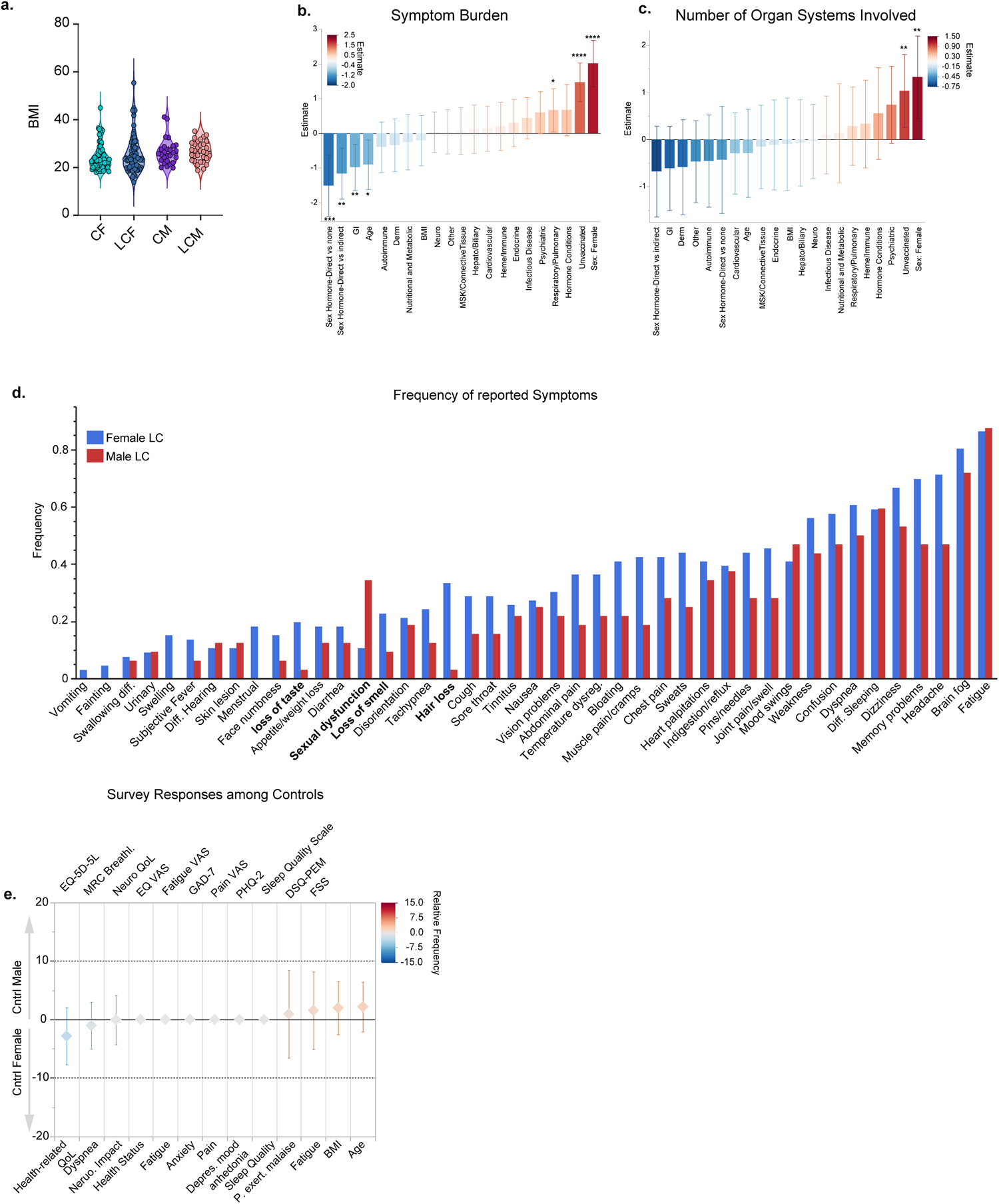
Sex differences in long COVID symptoms. (a) Comparison of BMI across sex-based groups. Groups were compared using Ordinary Least Squares Regression with ANOVA with an interaction term for sex and disease status. (b, c) Poisson GLMs with loglink MLE comparing the association of symptom burden with comorbidities, and other grouping classifications including vaccination status, and hormone therapies (b) and organ system involvement (c) amongst females and males with LC. Centered and scaled estimates are shown with negative values being negative predictors and positive values being positive predictors. Included parameters are labeled on the x-axis. Whiskers represent the 95% CI of the estimate value. P-values are adjusted for the simultaneous inclusion of multiple predictors in the model. Significant values are depicted by asterisks as follows *=p<.05, ** <.01, *** <.001, **** <.0001. (d) Frequency of reported symptoms by sex amongst individuals with LC. Bar graphs show the proportion of females and males reporting experiencing each symptom. Symptoms are ordered by increasing frequency from left to right. (f) Binomial Lasso regression with 5-fold cross validation comparing the relative frequencies of standardized health surveys across sex amongst control individuals while accounting for age and BMI. Whiskers represent the 95% confidence intervals.

**Extended Data Figure 2.**
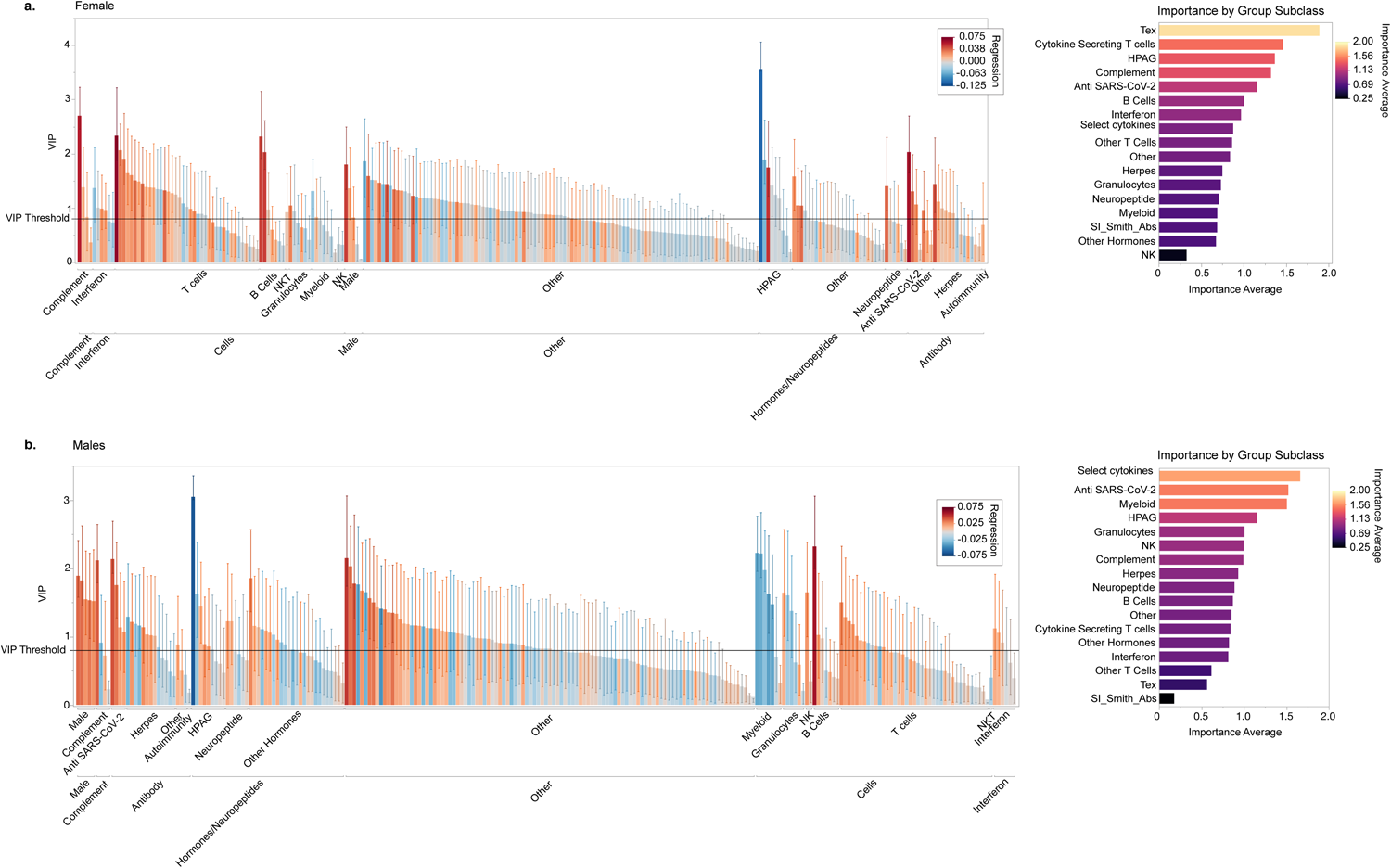
Sex-specific immune signatures in long COVID. (a., b. left) Bar plot depicting the variable of importance projection (VIP) score of each feature for prediction of Long COVID status versus controls amongst vaccinated individuals, without pre-existing autoimmune or hormone conditions or sex-hormone therapy in females (a) and males (b) with Long COVID versus their control counterparts in a sex-stratified manner. VIP measures were obtained from PLS-DA analysis with 5-fold cross-validation. Confidence intervals for each feature show bootstrapped 95% CI of the VIP score. Bar colors show regression value for each feature with positive predictors being more red and negative predictors being bluer as indicated by the legend. The VIP threshold cutoff of importance (0.8) is shown as a solid line. Features with 95% confidence intervals above the threshold cutoff of 0.8 and whose regression coefficient did not pass zero, were considered most significant and important. Detailed reports for each analysis can be found in supplemental data. Features were grouped by feature class. (a. b, right panels) and the average importance for each feature class is shown. Feature class is ordered by importance with the most important features on top. Color scheme also shows importance. For additional details on each PLS-DA model, please see supplementary Table X.

**Extended Data Figure 3.**
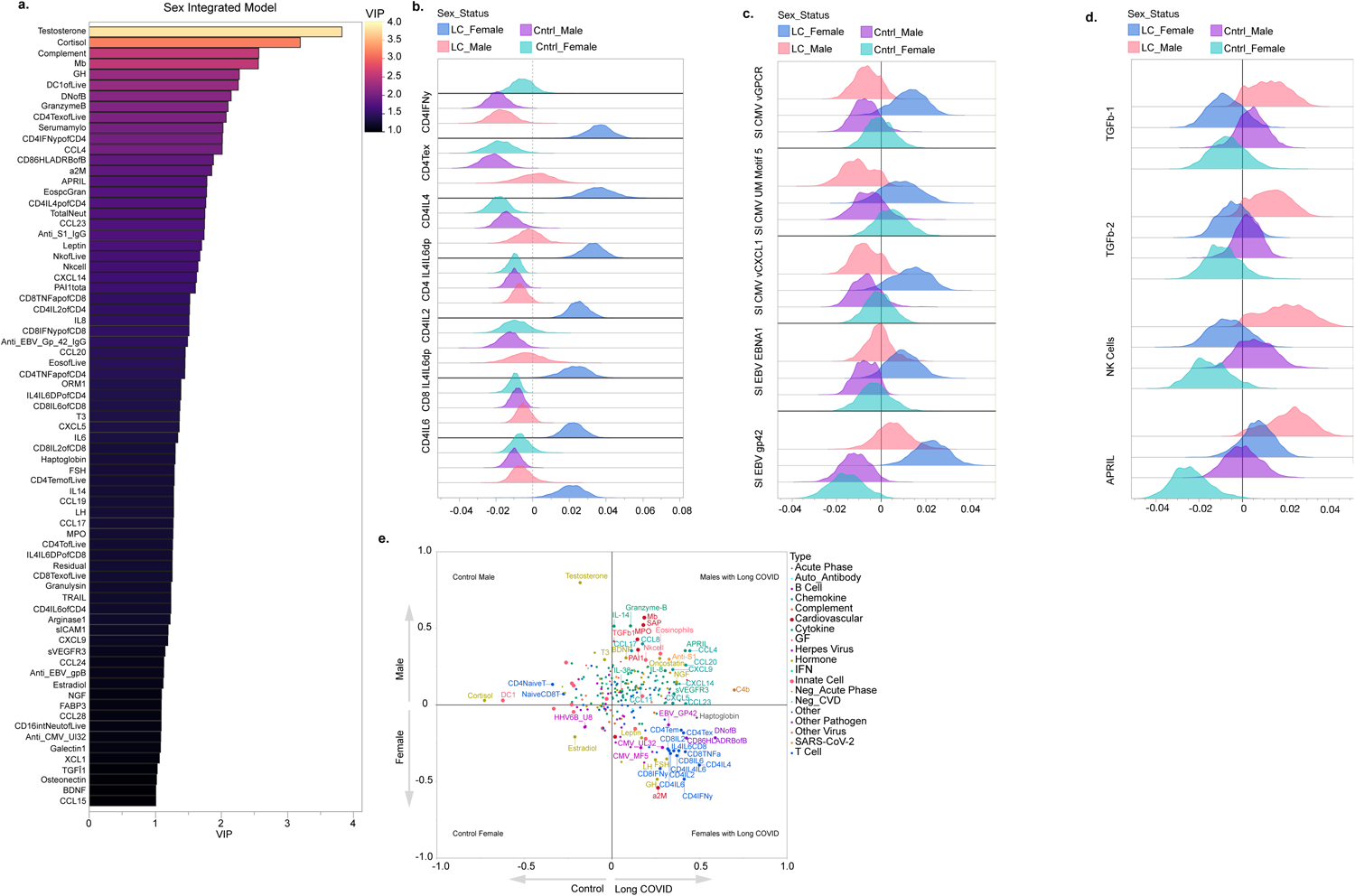
Sex-specific phenotypes identified by sex-integrated analysis. (a.) Bar plot depicting the features with the highest-ranking VIP scores for prediction of sex-based Long COVID status in a sex-integrated analysis amongst vaccinated individuals, without pre-existing autoimmune or hormone conditions or sex-hormone therapy. VIP measures were obtained from PLS-DA analysis with 5-fold cross-validation. Only features with bootstrapped 95%CI above the threshold cutoff of 0.8 are shown. Features are ranked by importance with the highest features on top. Color scheme denotes feature importance. (b-c.) Kernel density plots depicting the bootstrapped estimates for the relative expression of select T cells (a), herpesvirus epitopes (b), and cytokines (c) amongst all sex-based groups derived from the sex-integrated PLS-DA analysis as in a. (e) Feature coordinate plot demonstrating the relative expression of each feature amongst all four sex-based groups in a two-dimensional space. Sex based group classification is identified by the dotted line within each quadrant. Angular distance between a given feature and each sex-based group classification vector denotes the relative correlation of that feature with the specified sex-based group.

**Extended Data Figure 4.**
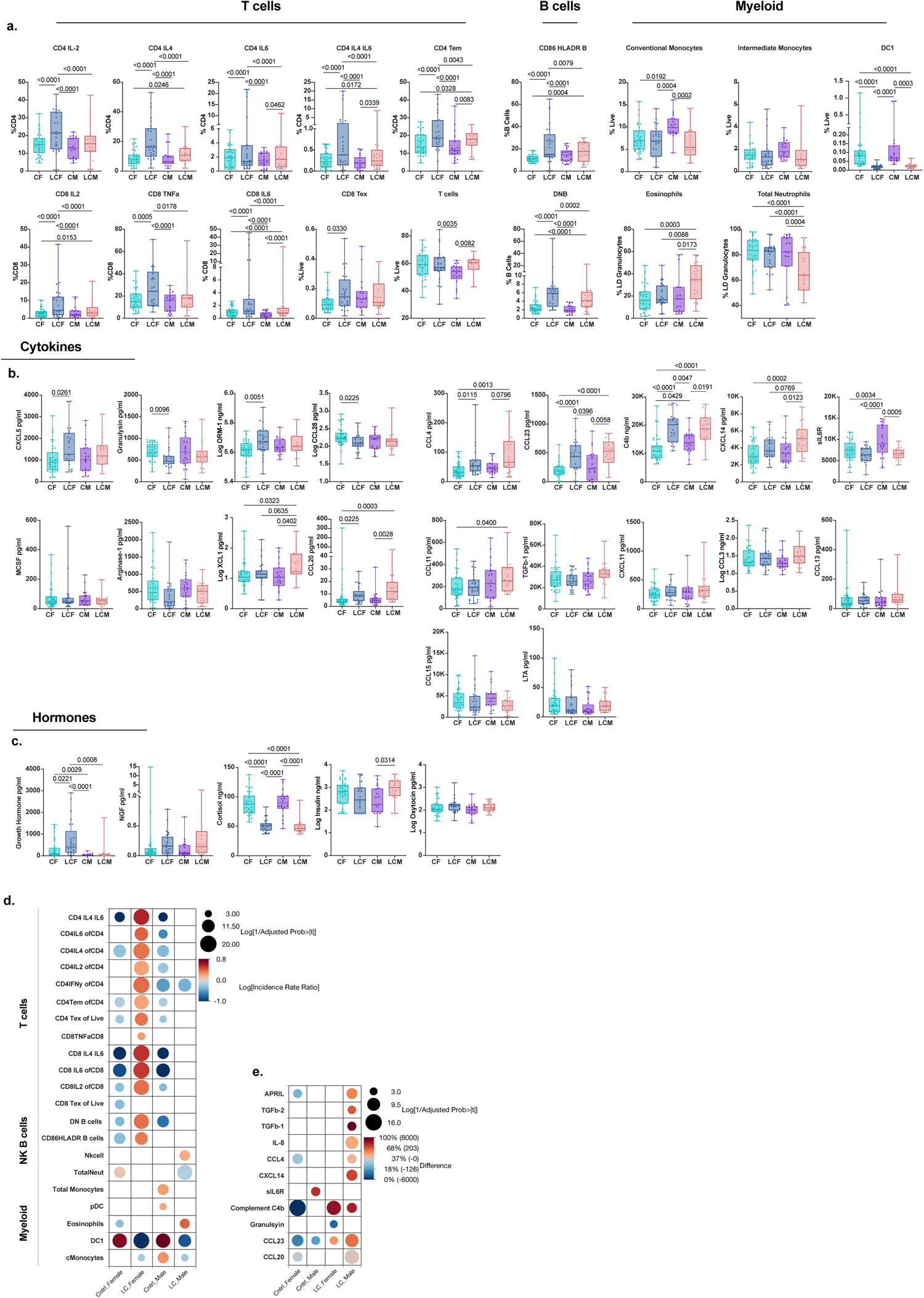
Sex-integrated analysis of top PLS-DA features associated with Long COVID status. (a) Generalized Linear Models (GLMs) with maximum likelihood estimation (MLE), comparing the frequency of T cell types, B cells, and Myeloid cells identified as predictors of LC status from PLS-DA analysis amongst vaccinated individuals across sex-based groups. Group inclusion is as in Figure 2. For frequencies less than 1 beta distribution was used with a logit model link transformation. For frequencies greater than 1 Poisson or negative binomial distribution was used with a Log Link transformation. Significance was derived from the estimate of each factor in the model. (b,c) Comparison of the top-predicted PLS-DA cytokine features (b) or hormones (c) associated with LC status across sex based groups. Comparisons across subgroups were conducted using mean estimates for normally distributed data or for data that conformed to normality following logarithmic transformation. Analysis of Variance (ANOVA) was applied, with Tukey’s adjustment for multiple comparisons to determine significance. For non-normal distributions, Kruskal-Wallis test was employed for initial assessment of significant differences across groups, followed by Steel-Dwass adjustment for multiple comparisons. (d) heatmap depicting relative enrichment of cell types across sex-based groups after adjustment for age and BMI. Values for the heatmap are derived from GLMs with Poisson distributions comparing the incidence rate ratio for each cell type frequency within each sex-based group relative the overall average using Analysis of Means (ANOM) with significance adjustment using the Nelson method. Comparisons were done while accounting for age and BMI. Color scheme denotes the relative incidence rate with red values showing a higher relative frequency over the average and blue values showing a lower relative frequency over the average. Only significant values are shown, and size of the circles are inversely proportional to the Log of the p-value. (e) Heatmap depicting the Analysis of Medians (ANOMed)s derived from GLMs using quantile regression to compare the median distribution of concentrations across sex-based groups to the overall median. Heatmap follows the same scheme as in d. Each cytokine was adjusted for age and BMI.

**Extended Data Figure 5.**
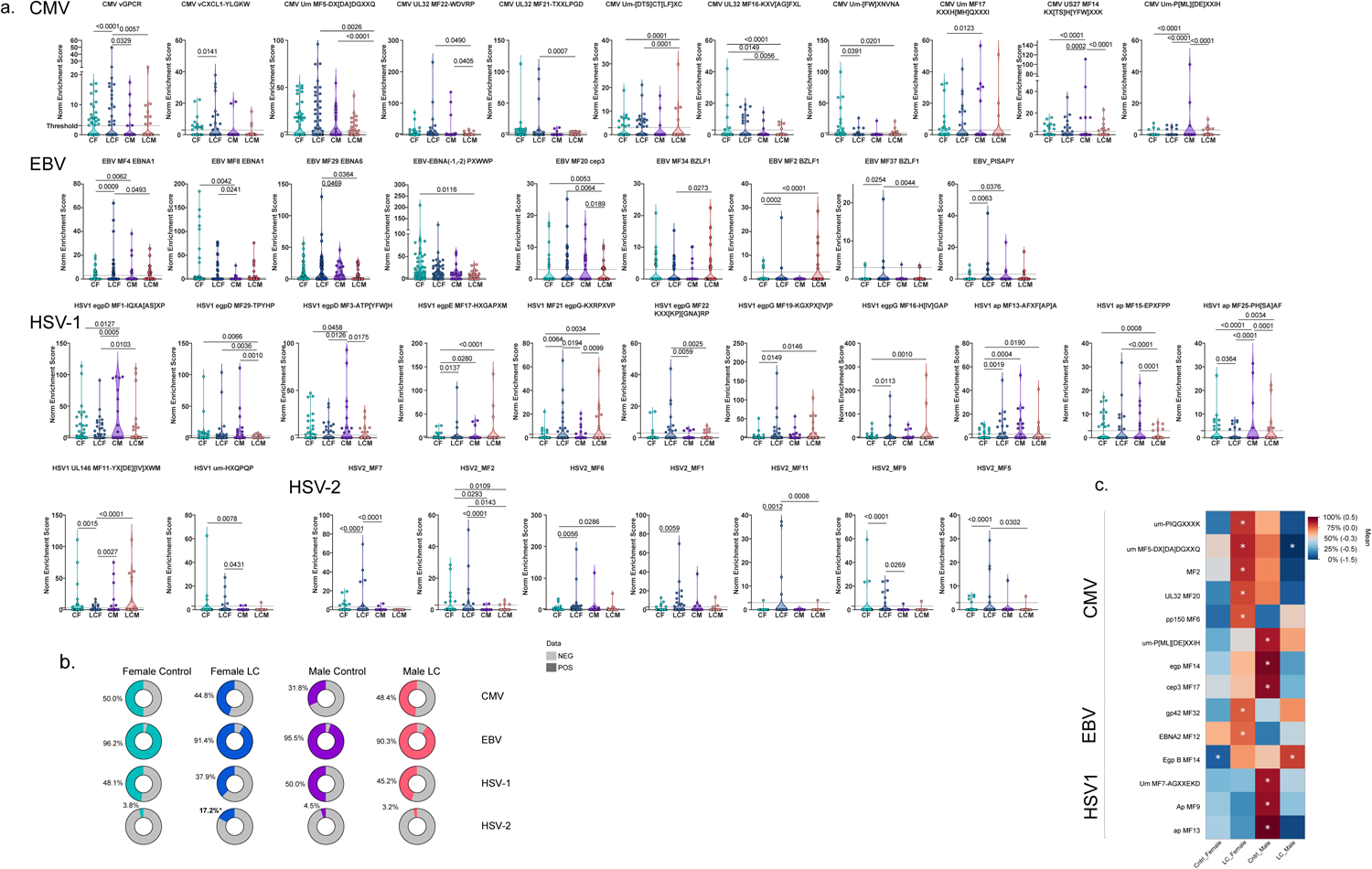
Sex differences in antibodies against viruses. (a). Comparison of SERA Motif normalized enrichment for specific motifs designated for CMV, EBV, HSV-1, and HSV-2 across sex-based groups using Generalized Linear Models (GLMs) with Zero inflated (ZI) negative binomial, or ZI Poisson maximum likelihood estimation (MLE) and loglink transformation. Significance was derived from the estimate of each factor in the model. (b.) Depiction and comparison of inferred positivity for CMV, EBV, HSV-1 and HSV-2 across sex-based groups relative to the overall average. Comparisons using Generalized Binomial regression for prediction of inferred positivity and comparisons were maded using the Analysis of Means (ANOM) comparing the frequency of positivity within each sex-based group to the overall average. Percent of inferred positive indivdiuals across each group is shown and colored as indicated in the legend. Significant values are bolded and denoted by an asterisk. C) Heatmap depicting the Analysis of Means of Transformed Ranks (ANOMTR) amongst inferred positive individuals for each sex based group. Comparsions were made amongst vaccinated individuals, without pre-existing autoimmune, hormone conditions or sex-hormone therapy as in Figure 2 and only amongst those who were inferred positive by SERA analysis for each herpesvirus shown ( for CMV: [CF=20, LCF=14, LCM=9 CM=7]; for EBV [CF=38, LCF=26, LCM=20, CM=20]; for HSV-1 [CF=17, LCF=10, CM=10, LCM=7]). Only significantly enriched motifs are shown. Significance adjustment was calculated using the Nelson method. Asterisks denote significance as p<.05.

**Extended Data Figure 6.**
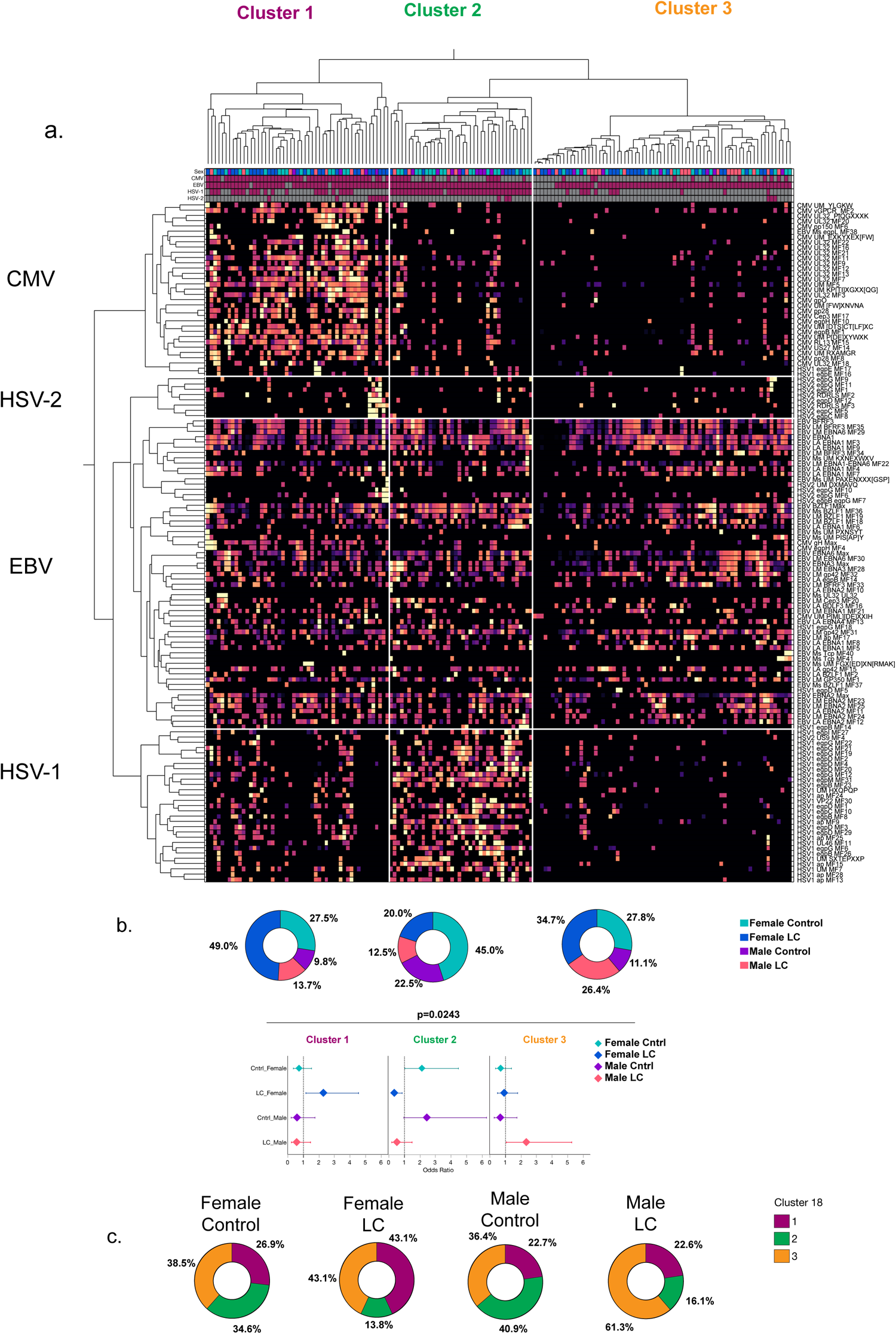
Unsupervised hierarchical clustering of herpesvirus reactivities within the MYLC cohort. (a) Unsupervised hierarchical clustering of normalized enrichment scores to linear epitopes associated with several herpesvirus reactivities including HSV-1, EBV, HSV-2, and CMV across the MYLC cohort using 2-way Ward based clustering. Standardization is column-based. Heatmap denotes normalized enrichment with brighter colors indicating higher relative normalized enrichment and darker colors indicating lower normalized enrichment. For the linear motif scores, four distinct clusters were identified and coincided with strong predominance of linear motifs designated to one of the herpesviruses. These four clusters were named for their herpesvirus dominance and are labeled above appropriately. In addition, three participant-based clusters were derived, labeled Cluster 1-3 as shown. Left heatmap panels (left to right) show the labels for each participant color-coded based on their sex-disease group designation as indicated by the figure key. Subsequent panels show the inferred positivity of each participant for each herpesvirus as indicated on the bottom of the heatmap, with maroon colors indicating inferred positivity. (b) Relative sex-based group composition for each cluster. Comparison of relative enrichment of each sex-based group across the three clusters was done using multinomial logistic regression. P-value shows the significance of sex-based group composition as a predictor of cluster composition. (c) Proportional composition of each sex-based group across the three clusters. Comparison for significance within each cluster was done in a sex-stratified manner using nominal logistic regression for prediction of disease status across each cluster. Significant enrichment of disease status for each cluster was compared to the overall average composition across clusters using ANOM. Significant percent enrichments are bolded and denoted by an asterisk.

**Extended Data Figure 7.**
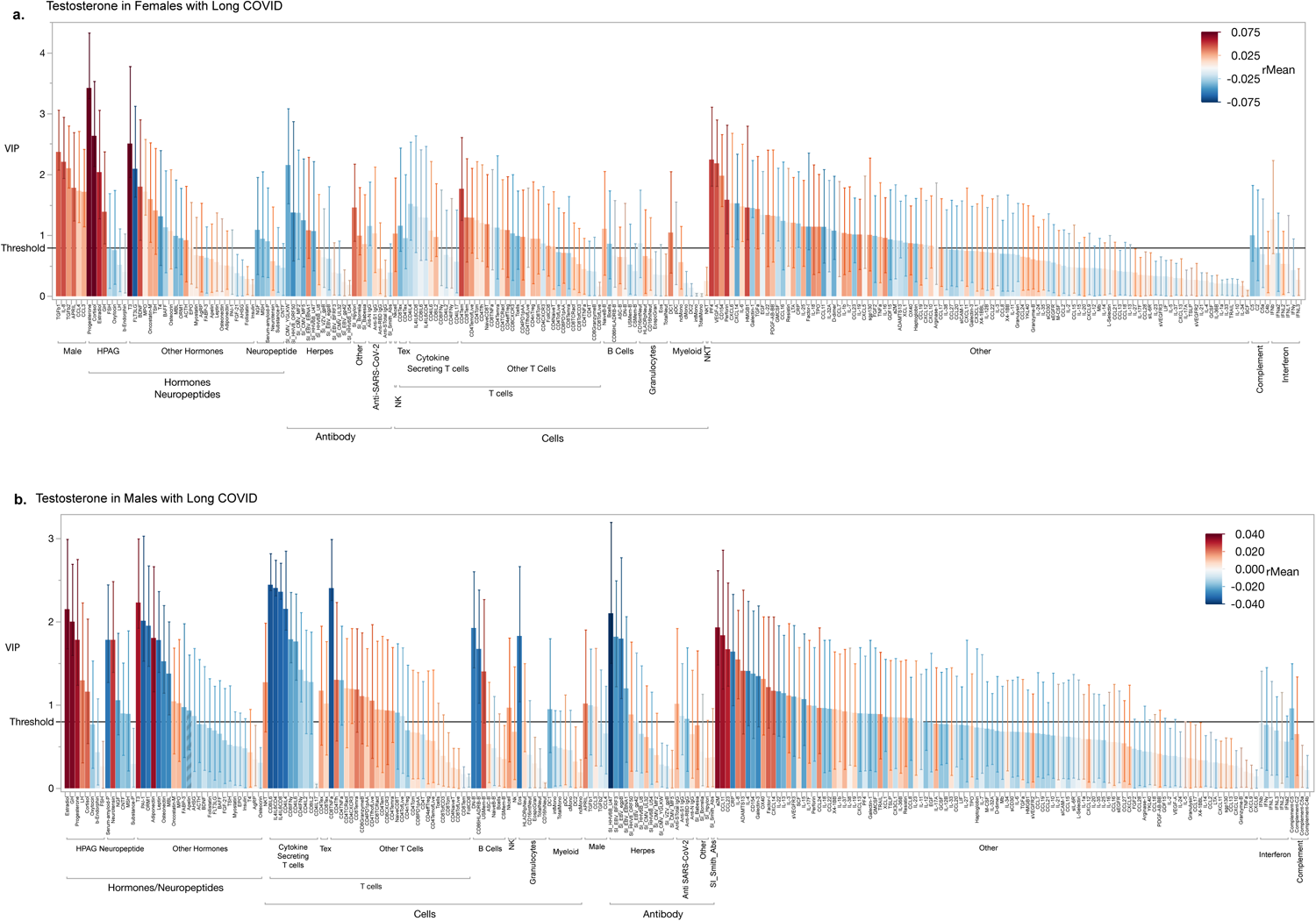
Predictors of testosterone levels in females and males with LC. (a., b.) Bar plot depicting the variable of importance projection (VIP) score of each feature for prediction of testosterone levels amongst individuals without pre-existing autoimmune or hormone conditions or sex-hormone therapy in females (a) and males with Long COVID. VIP measures were obtained from NIPALS analysis with k-fold (k=5) cross-validation. Bootstrapped 95% CI for the VIP score of each feature is shown. Bar colors show regression values for each feature with positive predictors being redder and negative predictors being bluer as indicated by the legend. The VIP threshold cutoff of importance (0.8) is shown as a solid line. Features with 95% confidence intervals above the threshold cutoff of 0.8 and whose regression coefficient did not pass zero, were considered most significant and important. Features were grouped by feature class.

**Extended Data Figure 8.**
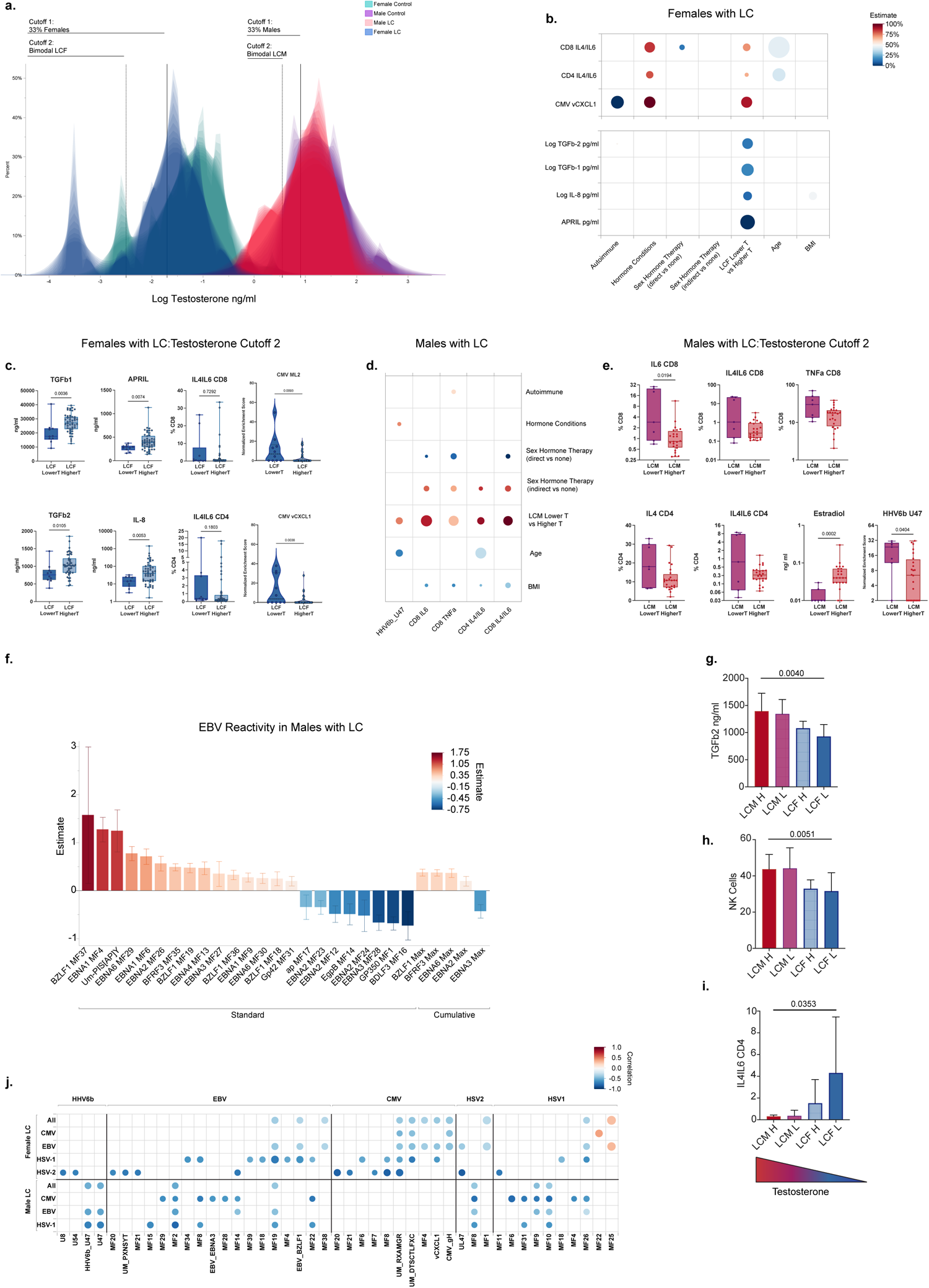
Immune signatures associated with testosterone levels. (a.) Shadowgram depicting the distribution of testosterone levels across females with LC and control females. Solid line denotes the division between the top two terciles and the bottom tercile accounting for approximately 66.6% and 33.3% of the distribution respectively. (b) Heatmaps showing the adjusted covariate effect of Lower Testosterone (LowerT) amongst females with LC for cell populations, linear epitopes, and cytokines shown in Figure 4b, Effect estimates for each factor was adjusted by inclusion of each covariate in the model including age, BMI, presence or absence of autoimmunity, hormone conditions, or hormone therapies. Estimates for cell populations were done using Poisson regression and for epitope normalized enrichment using a zero-inflated Poisson regression model. Effect estimates for cytokines were derived from a linear GLM after Log transformation for non-normal values. Effects for each factor are shown on a color scale with effects positively associated with LowerT in females with LC having a red color distribution and blue for negative association with LowerT in females with LC. Size of the circles denote the relative significance of each cofactor within each model. Only significant circles are shown. (c) Shadowgram depicting the distribution of testosterone levels across males with LC and control males. Solid line denotes the division between the top two terciles and the bottom tercile accounting for approximately 66.6% and 33.3% of the distribution respectively. (d) Heatmaps showing the adjusted covariate effect of Lower Testosterone (LowerT) amongst males with LC for cell populations, and linear epitopes, shown in Figure 4d Heatmap was constructed as in (b). (e) Comparison of linear epitope reactivity amongst males with Long COVID in lower testosterone (LowerT) and higher testosterone (HigherT) groups. Comparisons to each linear motif were done using generalized linear mixed models (GLMMs) with Poisson distribution. individual variability and repeated measures were accounted for as a random effect in the model. Only linear peptides for which at least two individual males with LC and at least one individual in each testosterone group had a non-zero value were compared. Data was done on individual motifs and by protein maximum. Significance was adjusted using Benjamini FDR correction. (f-h) Comparison of linear trends amongst sex and testosterone-based groupings of females and males with LC for key identified factors including TGF-β12 (f), NK cells (g), and IL4/6 double positive CD4 T cells (h). (i) Heatmap of bootstrapped Pearson correlations of testosterone levels and herpesvirus reactivities amongst females and males with LC. Males and females were analyzed separately and stratified by presumptive positivity for either EBV, or CMV based on SERA classifications (EBV Pos= Presumptive EBV positive; CMV Pos= presumptive CMV positive; All= all females or all males with Long COVID).

**Extended Data Table 1:**
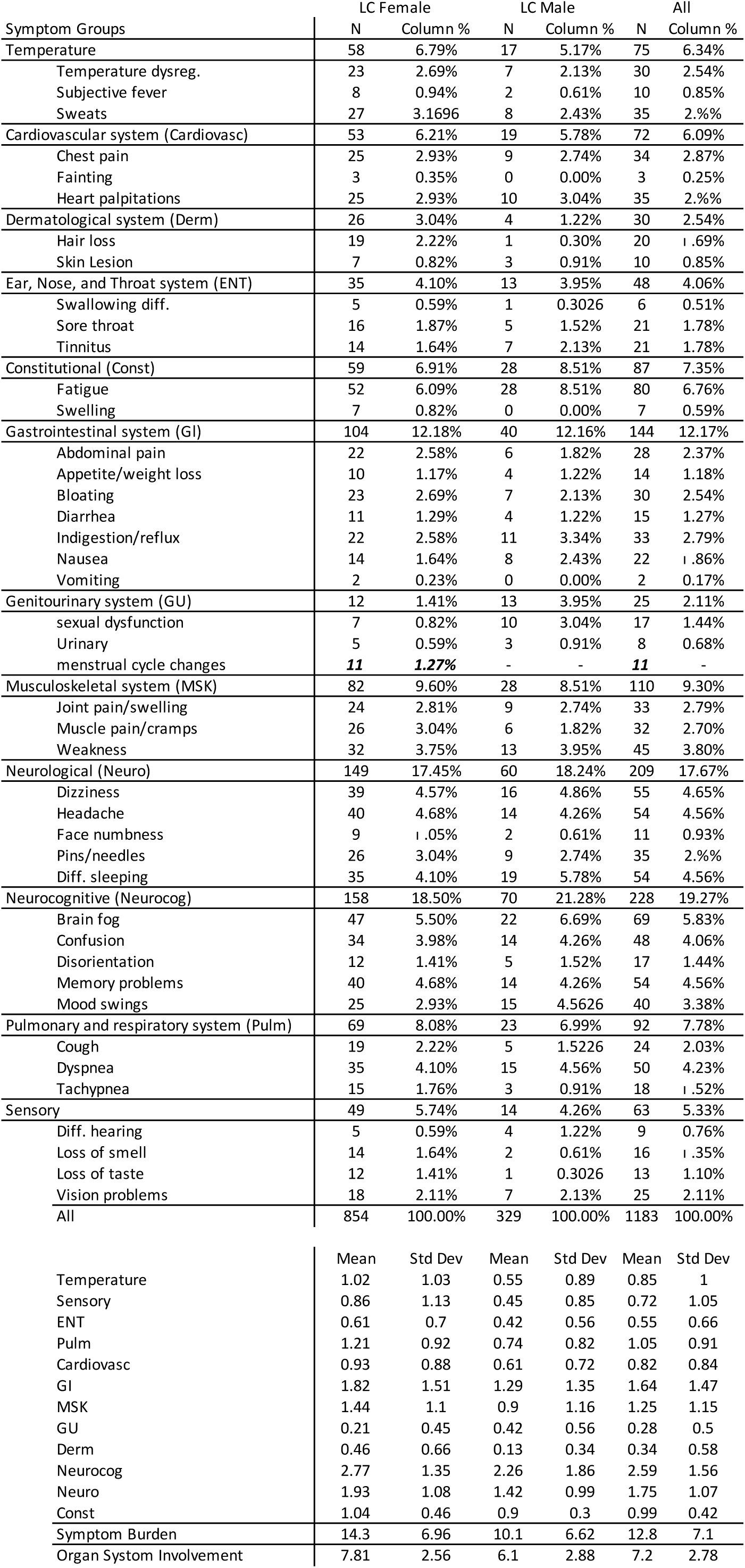
Clinical Demographics of MY-LC Cohort. Summary of demographic and clinical characteristics for the MY-LC Study. Participants were stratified into four sex-based groups: 185 participants were initially enrolled at Mount Sinai Hospital, in New York City, New York, composed of 101 individuals with LC (69 females, 32 males), and 82 control subjects (58 females, 24 males)^8^. Post-enrollment, a comprehensive review of electronic medical records led to the identification of 20 individuals with potential confounding factors: outlier conditions (n=7), oral steroid use (n=4), mismatched data (n=3), and missing data (n=6). To uphold the integrity of our primary analysis, these 20 individuals were excluded from the primary dataset, leading to a refined cohort of 165 participants. Various demographic features and characteristics are reported by row for each cohort. Within each cell, Counts or feature averages are reported with 95% Cis where needed, Statistical tests are reported as p-values and accompanying test statistics:

| Symptom Groups | LC Female |  | LC Male |  | All |  |
| --- | --- | --- | --- | --- | --- | --- |
|  | N | Column % | N | Column % | N | Column % |
| Temperature | 58 | 6.79% | 17 | 5.17% | 75 | 6.34% |
| Temperature dysreg. | 23 | 2.69% | 7 | 2.13% | 30 | 2.54% |
| Subjective fever | 8 | 0.94% | 2 | 0.61% | 10 | 0.85% |
| Sweats | 27 | 3.1696 | 8 | 2.43% | 35 | 2.9% |
| Cardiovascular system (Cardiovasc) | 53 | 6.21% | 19 | 5.78% | 72 | 6.09% |
| Chest pain | 25 | 2.93% | 9 | 2.74% | 34 | 2.87% |
| Fainting | 3 | 0.35% | 0 | 0.00% | 3 | 0.25% |
| Heart palpitations | 25 | 2.93% | 10 | 3.04% | 35 | 2.9% |
| Dermatological system (Derm) | 26 | 3.04% | 4 | 1.22% | 30 | 2.54% |
| Hair loss | 19 | 2.22% | 1 | 0.30% | 20 | 1.69% |
| Skin Lesion | 7 | 0.82% | 3 | 0.91% | 10 | 0.85% |
| Ear, Nose, and Throat system (ENT) | 35 | 4.10% | 13 | 3.95% | 48 | 4.06% |
| Swallowing diff. | 5 | 0.59% | 1 | 0.3026 | 6 | 0.51% |
| Sore throat | 16 | 1.87% | 5 | 1.52% | 21 | 1.78% |
| Tinnitus | 14 | 1.64% | 7 | 2.13% | 21 | 1.78% |
| Constitutional (Const) | 59 | 6.91% | 28 | 8.51% | 87 | 7.35% |
| Fatigue | 52 | 6.09% | 28 | 8.51% | 80 | 6.76% |
| Swelling | 7 | 0.82% | 0 | 0.00% | 7 | 0.59% |
| Gastrointestinal system (GI) | 104 | 12.18% | 40 | 12.16% | 144 | 12.17% |
| Abdominal pain | 22 | 2.58% | 6 | 1.82% | 28 | 2.37% |
| Appetite/weight loss | 10 | 1.17% | 4 | 1.22% | 14 | 1.18% |
| Bloating | 23 | 2.69% | 7 | 2.13% | 30 | 2.54% |
| Diarrhea | 11 | 1.29% | 4 | 1.22% | 15 | 1.27% |
| Indigestion/reflux | 22 | 2.58% | 11 | 3.34% | 33 | 2.79% |
| Nausea | 14 | 1.64% | 8 | 2.43% | 22 | 1.86% |
| Vomiting | 2 | 0.23% | 0 | 0.00% | 2 | 0.17% |
| Genitourinary system (GU) | 12 | 1.41% | 13 | 3.95% | 25 | 2.11% |
| sexual dysfunction | 7 | 0.82% | 10 | 3.04% | 17 | 1.44% |
| Urinary | 5 | 0.59% | 3 | 0.91% | 8 | 0.68% |
| menstrual cycle changes | **11** | **1.27%** | - | - | **11** | - |
| Musculoskeletal system (MSK) | 82 | 9.60% | 28 | 8.51% | 110 | 9.30% |
| Joint pain/swelling | 24 | 2.81% | 9 | 2.74% | 33 | 2.79% |
| Muscle pain/cramps | 26 | 3.04% | 6 | 1.82% | 32 | 2.70% |
| Weakness | 32 | 3.75% | 13 | 3.95% | 45 | 3.80% |
| Neurological (Neuro) | 149 | 17.45% | 60 | 18.24% | 209 | 17.67% |
| Dizziness | 39 | 4.57% | 16 | 4.86% | 55 | 4.65% |
| Headache | 40 | 4.68% | 14 | 4.26% | 54 | 4.56% |
| Face numbness | 9 | 1.05% | 2 | 0.61% | 11 | 0.93% |
| Pins/needles | 26 | 3.04% | 9 | 2.74% | 35 | 2.9% |
| Diff. sleeping | 35 | 4.10% | 19 | 5.78% | 54 | 4.56% |
| Neurocognitive (Neurocog) | 158 | 18.50% | 70 | 21.28% | 228 | 19.27% |
| Brain fog | 47 | 5.50% | 22 | 6.69% | 69 | 5.83% |
| Confusion | 34 | 3.98% | 14 | 4.26% | 48 | 4.06% |
| Disorientation | 12 | 1.41% | 5 | 1.52% | 17 | 1.44% |
| Memory problems | 40 | 4.68% | 14 | 4.26% | 54 | 4.56% |
| Mood swings | 25 | 2.93% | 15 | 4.5626 | 40 | 3.38% |
| Pulmonary and respiratory system (Pulm) | 69 | 8.08% | 23 | 6.99% | 92 | 7.78% |
| Cough | 19 | 2.22% | 5 | 1.5226 | 24 | 2.03% |
| Dyspnea | 35 | 4.10% | 15 | 4.56% | 50 | 4.23% |
| Tachypnea | 15 | 1.76% | 3 | 0.91% | 18 | 1.52% |
| Sensory | 49 | 5.74% | 14 | 4.26% | 63 | 5.33% |
| Diff. hearing | 5 | 0.59% | 4 | 1.22% | 9 | 0.76% |
| Loss of smell | 14 | 1.64% | 2 | 0.61% | 16 | 1.35% |
| Loss of taste | 12 | 1.41% | 1 | 0.3026 | 13 | 1.10% |
| Vision problems | 18 | 2.11% | 7 | 2.13% | 25 | 2.11% |
| All | 854 | 100.00% | 329 | 100.00% | 1183 | 100.00% |

|  | Mean | Std Dev | Mean | Std Dev | Mean | Std Dev |
| --- | --- | --- | --- | --- | --- | --- |
| Temperature | 1.02 | 1.03 | 0.55 | 0.89 | 0.85 | 1 |
| Sensory | 0.86 | 1.13 | 0.45 | 0.85 | 0.72 | 1.05 |
| ENT | 0.61 | 0.7 | 0.42 | 0.56 | 0.55 | 0.66 |
| Pulm | 1.21 | 0.92 | 0.74 | 0.82 | 1.05 | 0.91 |
| Cardiovasc | 0.93 | 0.88 | 0.61 | 0.72 | 0.82 | 0.84 |
| GI | 1.82 | 1.51 | 1.29 | 1.35 | 1.64 | 1.47 |
| MSK | 1.44 | 1.1 | 0.9 | 1.16 | 1.25 | 1.15 |
| GU | 0.21 | 0.45 | 0.42 | 0.56 | 0.28 | 0.5 |
| Derm | 0.46 | 0.66 | 0.13 | 0.34 | 0.34 | 0.58 |
| Neurocog | 2.77 | 1.35 | 2.26 | 1.86 | 2.59 | 1.56 |
| Neuro | 1.93 | 1.08 | 1.42 | 0.99 | 1.75 | 1.07 |
| Const | 1.04 | 0.46 | 0.9 | 0.3 | 0.99 | 0.42 |
| Symptom Burden | 14.3 | 6.96 | 10.1 | 6.62 | 12.8 | 7.1 |
| Organ System Involvement | 7.81 | 2.56 | 6.1 | 2.88 | 7.2 | 2.78 |

**Extended Data Table 2:** Sex-stratified analysis of past medical history between individuals with Long COVID and their control counterparts. Each feature was analyzed independently using multivariate nominal logistic regression which accounted for age and BMI.

Sex-Stratified Comorbidities
| Past Medical History | Female |  |  |  |  | Male |  |  |  |  |
| --- | --- | --- | --- | --- | --- | --- | --- | --- | --- | --- |
|  | Odds Ratio | Lower CI | Upper CI | Prob>ChiSq | FDR Adj PValue | Odds Ratio | Lower CI | Upper CI | Prob>ChiSq | FDR Adj PValue |
| Previously identified Asthma | 4.66779549 | 1.2912117 | 16.8743164 | 0.0188 | - | 3.55461893 | 0.66547815 | 18.986823 | 0.1379 | - |
| **Past Medical History** |  |  |  |  |  |  |  |  |  |  |
| Autoimmune | 0.88927199 | 0.20069382 | 3.94035399 | 0.87720823 | 0.935688777 | - | - | - | - | - |
| Cardiovascular | 4.2726141 | 1.35724595 | 13.4502012 | 0.01306372 | 0.052254869 | 2.52106395 | 0.71211369 | 8.92520887 | 0.15168956 | 0.599123859 |
| Derm | 1.22531796 | 0.24127226 | 6.22286245 | 0.80639299 | 0.921591994 | 0.283816 | 0.02305702 | 3.49357922 | 0.32545926 | 0.616367027 |
| Endocrine: Direct Hormone Conditions | 1.99387356 | 0.71365922 | 5.57063048 | 0.18803232 | 0.342241727 | 3.1375824 | 0.54652678 | 18.0127008 | 0.19970795 | 0.599123859 |
| Endocrine: Nutritional/Metabolic/Other Endocrine | 0.27127003 | 0.05258142 | 1.39949487 | 0.11912392 | 0.272283242 | 0.82069305 | 0.0791289 | 8.5118979 | 0.86848834 | 0.998468575 |
| Genitourinary | 8.71125505 | 0.93278011 | 81.354613 | 0.05757506 | 0.153533489 |  | 0 |  | 0.99825111 | 0.998468575 |
| GI | 8.62335812 | 2.16736057 | 34.3100757 | 0.00222979 | 0.017838355 | 7.19580564 | 1.19481828 | 43.3368151 | 0.03121967 | 0.234147517 |
| Heme/Onc/Immun | 5.67047849 | 1.08801234 | 29.5532736 | 0.03938833 | 0.126042669 | 0.29261992 | 0.02218318 | 3.85997104 | 0.35044965 | 0.616367027 |
| Hepato/Biliary | 186292607 | 0 |  | 0.99776441 | 0.997764412 | 3.40594442 | 0.34701863 | 33.4289177 | 0.29293664 | 0.616367027 |
| Infectious Disease | 1.25553724 | 0.30248406 | 5.21142752 | 0.75399961 | 0.921591994 | 0.56960276 | 0.03201195 | 10.1351945 | 0.7015891 | 0.956712412 |
| MSK/ConnectiveTissue | 3.06838572 | 0.56446185 | 16.6795879 | 0.19431605 | 0.342241727 | 2.8583414 | 0.28792819 | 28.3755318 | 0.36982022 | 0.616367027 |
| Neuro | 1.98329283 | 0.67364774 | 5.83903167 | 0.21390108 | 0.342241727 | 686092519 | 0 |  | 0.99846858 | 0.998468575 |
| Psychiatric | 0.87339717 | 0.34421783 | 2.21610432 | 0.77569162 | 0.921591994 | 1.65842066 | 0.49541505 | 5.55162602 | 0.41186986 | 0.617804783 |
| Respiratory/Pulmonary | 4.90524185 | 1.39605937 | 17.2352251 | 0.01017987 | 0.052254869 | 2.4406159 | 0.64846964 | 9.18563585 | 0.18702069 | 0.599123859 |
| None | 0.09232516 | 0.03410367 | 0.24994185 | 2.75E-06 | 4.39994E-05 | 0.19157188 | 0.05254049 | 0.69850489 | 0.01229449 | 0.184417404 |
| Other | 1.55888436 | 0.60791501 | 3.99746745 | 0.35546205 | 0.517035709 | 0.82216325 | 0.18939583 | 3.56899302 | 0.79376745 | 0.992209317 |
| **Sex Hormone Medications** |  |  |  |  |  |  |  |  |  |  |
| Direct | 3.11616777 | 0.92481469 | 10.499943 | 0.0667 | - | - | - | - | - | - |
| Indirect | - | - | - | - | - | 1.52194764 | 0.12626329 | 18.3451947 | 0.7409 | - |
| **Vaccination Status** |  |  |  |  |  |  |  |  |  |  |
| Unvaccinated | 11.7942061 | 1.36810277 | 101.676059 | 0.0248 | - | - | - | - | - | - |

**Extended Data Table 3:**
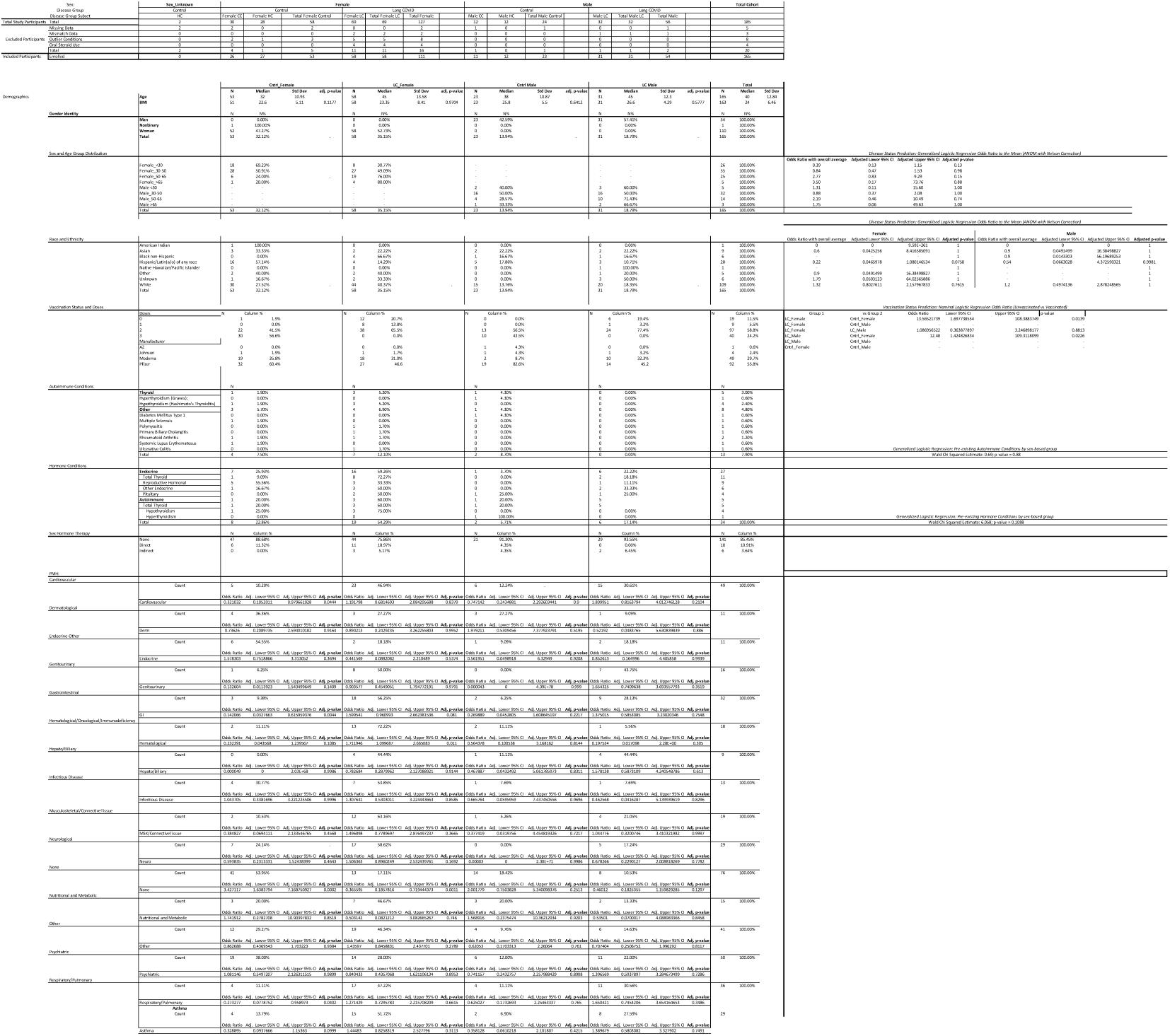
Organ System Grouping Composition. Table shows the inclusion of each symptom into organ systems and the frequency of each symptom across sex-based groups. For direct comparisons across sex, symptoms that were sex-specific, such as changes in menstrual cycle, were excluded but were incorporated in all sex-intrinsic comparisons.

